# Discovery and validation of serum metabolic signature of neonatal sepsis

**DOI:** 10.1101/2024.03.07.24303587

**Authors:** Riya Ahmed, Suraj Thottathil, Nidhi Yadav, Anil Behera, Adyasha Sarangi, Vrinda Satheesan Nair, Pradeep Debata, Rajni Gaind, Gita Prashad Kaushal, Renu Gur, Ravi Sachan, Kirti Nirmal, Ravinder Kaur, Sushma Nangia, Vivek Kumar, Nupur Sharma, Jaswinder Singh Maras, Saroj Kant Mohapatra, Krishnamohan Atmakuri, Ramesh Agarwal, Mari Jeeva Sankar, Ranjan Kumar Nanda

## Abstract

A more accurate diagnostic biosignature is crucial for neonatal sepsis. In this report, we identified a serum metabolite signature for diagnosing neonatal sepsis cases using mass spectrometry-based profiling of serum samples from two discovery cohorts (set-I/-II: n=71/269) of sepsis patients (culture positive/negative: CP/CN) and controls (no-sepsis: NS or healthy controls: HC). This signature was validated in both cross-sectional (n=60) and follow-up cohorts (n=100). The six-metabolite signature, which includes 1,5-anhydro-D-sorbitol, lactic acid, malic acid, myo-inositol, phenylalanine, and lysine, can distinguish CP and CN sepsis cases from HC. The deregulated serum metabolites returned to HC levels in neonates after completing antibiotic treatment. Additionally, a metabolic signature of PE (20:4(5Z,8Z,11Z,14Z)/0:0), 12-amino-dodecanoic acid, and 1,5-anhydro-D-sorbitol identified from a validation set-II (n=100) using LC-MS could differentiate CP and CN groups from NS groups. Translating this serum metabolite signature into a simple, deployable blood test for neonatal sepsis could enable faster and more accurate decision-making.

## INTRODUCTION

Approximately 2 million sepsis cases are reported worldwide, with an 11-19% mortality rate.^1,2^ Sepsis is a life-threatening condition characterized by multi-organ dysfunction caused by a dysregulated host response to infection, often due to pathogens such as *Acinetobacter baumannii, Escherichia coli,* and *Klebsiella* spp.^3,4^ Neonatal sepsis may present differently from adult sepsis due to the immature immune systems and gut microbiota composition; early diagnosis is crucial for timely intervention and improving survival rates.^5^ Neonatal sepsis that occurs within 72 hours of birth is classified as early-onset sepsis (EOS), while cases presenting after this period are grouped as late-onset sepsis (LOS). EOS can be acquired in utero or during vaginal delivery, whereas LOS is typically linked to hospital-acquired infections. Culture-positive (CP) sepsis cases are associated with a higher risk of death, longer hospital stays, and the need for mechanical ventilation compared to neonates with culture-negative (CN) sepsis.^6^

Sepsis-induced aggressive immune responses may influence metabolic pathways in affected cells and tissues and could be reflected in their systemic circulation. Currently, serum procalcitonin (PCT) and C-reactive protein (CRP) are biomarkers used for the clinical diagnosis of sepsis but have low specificity.^6,7^ Other biomarkers like cluster of differentiation 64 (CD64), CD11b, interferon gamma-induced protein 10 (IP-10), interleukin 12 (IL-12), and soluble triggering receptor expressed on myeloid cell-1 (sTREM-1) show higher predictive ability in adult sepsis.^8^ In neonates, a combination of immature to total neutrophil ratio (I/T ratio), CRP levels, total blood count, and total leukocyte count-erythrocyte sedimentation rate (TLC ESR) is used for sepsis screening. Blood metabolites such as lactate and phenylalanine levels are reported to correlate directly with disease severity and mortality in adult sepsis shock patients; however, their combined predictive ability has not been reported.^8,9^ Mass spectrometry-based global biofluid metabolic profiling in sepsis patients and controls may identify key deregulated molecules to develop diagnostic biosignatures and improve understanding of the disease pathophysiology.^10^ For example, in an adult sepsis patient cohort, a set of nine metabolite signatures with high predictive capability were identified.^11^ In this study, serum metabolites from neonates with sepsis and two control groups were analyzed using gas or liquid chromatography-based mass spectrometry (GC/LC-MS) to identify a putative biosignature and serially monitored in neonates completing antibiotic therapy. Identifying and validating the sepsis serum molecular signature is essential for developing novel diagnostic tools to classify patients quickly, and a better understanding of the disrupted pathways in sepsis will aid in creating host-directed therapeutics.

## RESULTS

### Clinical details of the study subjects

The clinical details of the neonates (n=600, 41.3% female) with sepsis (CP: n=148, CN: n=168, Figure 1) and controls (no sepsis, NS: n=161; healthy controls, HC: n=123) included in this study are presented in Figure 2, Supplementary Tables S1 and S2. Most CP (92.3%) and CN (95.2%) neonates had EOS. In the discovery set-I, blood samples from 37.5% of CP cases tested positive for *Acinetobacter baumannii* and 19% for coagulase-negative Staphylococcus (CoNS), as shown in Supplementary Table S3. In discovery set-II, 20.6% of CP cases involved *Acinetobacter baumannii*, and 28.6% involved *Klebsiella pneumoniae*. *Acinetobacter baumannii* accounted for 33.3% of CP cases in validation set-I. In the follow-up set, the prevalence was 13.8% for *Acinetobacter baumannii* and 20.7% for *Klebsiella pneumoniae*. In the independent validation set-II used for LC-MS analysis, incidence rates were 12% for *Acinetobacter baumannii* and 16% for *Klebsiella pneumoniae*. The detailed counts of samples positive for each pathogen are provided in Supplementary Table S3.

**Figure 1:**
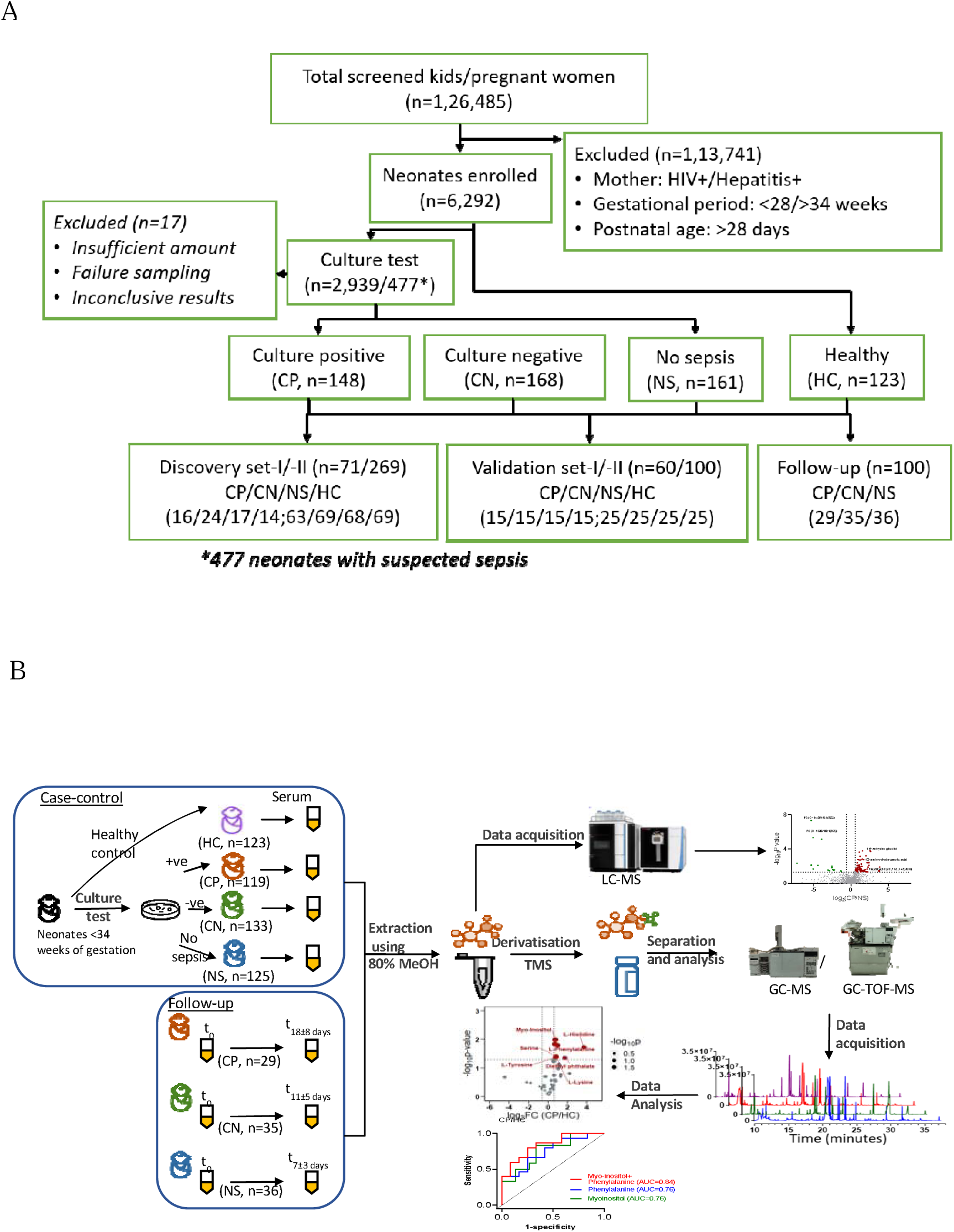
Overview of subject enrolment, grouping, distribution, and workflow used to identify and validate the serum markers for neonatal sepsis. (A) Screening and enrolment of suspected sepsis neonates and healthy controls, and their classification for serum metabolite signature identification and validation. (B) Blood culture was used to classify the subjects as case (culture positive: CP; culture negative: CN) and no sepsis (NS) group, which was used as a control along with healthy control (HC). Serum samples were used to extract metabolites for derivatization and mass spectrometry data acquisition and analysis using statistical tools. GC-MS: Gas Chromatography-Mass Spectrometry; GC-TOF-MS: Gas Chromatography-time-of-flight-Mass Spectrometry.

**Figure 2:**
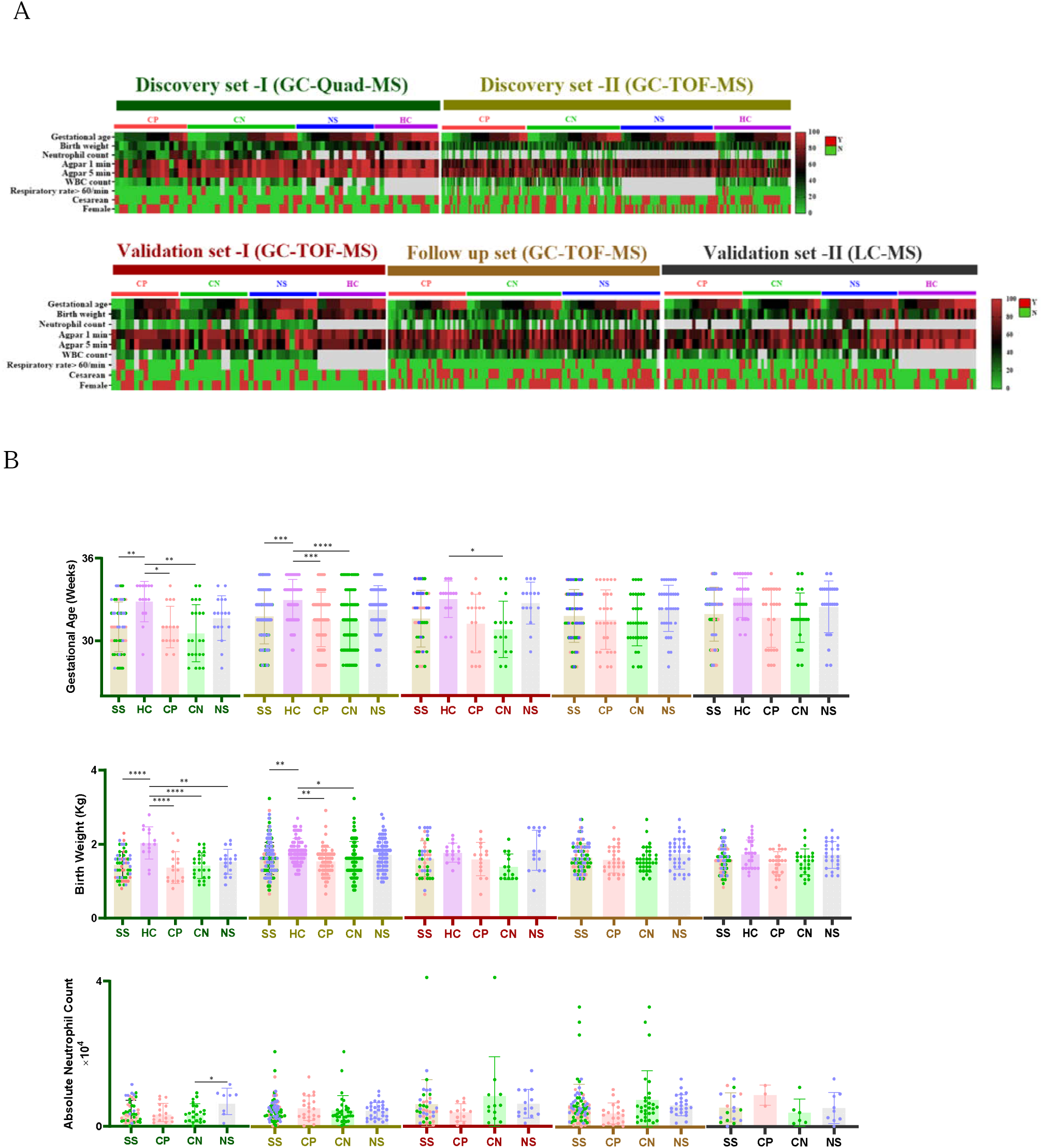
Epidemiological details of the study subjects and groups used in this study and the detailed one is presented in Supplementary Table S1. **A.** Clinical characteristics (gestational age, birth weight, caesarean deliveries, absolute neutrophil count) of the study participants used in the discovery sets I and II, validation sets I and II and the follow up set (Y/N:Yes/No) (B). Statistical comparisons between groups using One-way ANOVA with Tukey’s multiple comparison (P-value<0.05:*, <0.005:**, <0.0005:***, <0.0001:****).

The gestational age of neonates of CP (30.9±1.7 weeks, p=0.003) and CN (30.8±2.1 weeks, p=0.004) sepsis groups was significantly lower than that of the HC group (32.9±1.5 weeks) in the discovery set-I. The gestational age was also significantly lower for the sepsis groups compared to the HC and NS groups in the discovery set-II and validation set-I (Supplementary Table S4). In validation set-II, the CP, CN, and NS groups had lower gestational ages (CP/CN/NS: 31.1±1.9/31.1±1.5/31.8±1.6 weeks) than the HC groups (32.4±1.2 weeks, p<0.005). The gestational age distribution for each study group is shown in Supplementary Table S4. Similarly, the neonates of CP and CN groups had significantly lower birth weights (discovery set-I, CP/CN: 1.3±0.5/1.4±0.3 in kg; discovery set-II, CP/CN: 1.4±0.3/1.5±0.4 in kg) compared to their respective healthy controls (discovery set-I: 2.0±0.4 kg, p<0.0001; discovery set-II: 1.7±0.3 kg, p< 0.0003). The birth weight of neonates from validation set-I was lower in CN (1.3±0.3 kg) compared to HC (1.7±0.3 kg, p<0.004) and NS (1.7±0.5 kg, p<0.01) groups. In validation set-II, the CP and CN neonates had significantly lower birth weights (1.4±0.2/1.4±0.3 kg) than the NS (1.6±0.3 kg, p<0.08/0.03) and HC control groups (1.6±0.3 kg, p<0.008/0.03). In discovery set-I, the number of cesarean deliveries was lower in the CN group (23.8%) compared to the HC (43.8%). In discovery set-II, the number of cesarean deliveries was lower in CP (46.7%) than in the other groups. The CP groups in validation set-II had a higher rate of caesarean deliveries (48%), followed by NS (40%), then CN (32%), and healthy controls (28%). The absolute neutrophil count was significantly lower in the CP (3.8±2.5×10³) and CN (3.7±2.5×10³) groups compared to the NS (6.9±3.4×10³) group in discovery set-I. This difference was not significant in discovery set-II and validation set-I. In validation set-II, the absolute neutrophil count was higher (8.1±2.1×10^3^) in the CP group compared to CN (3.6±3.1×10^3^) and NS (4.8±3.7×10^3^) groups.

### Serum metabolic phenotyping of sepsis patients and controls of discovery set-I using mass spectrometry

Serum metabolite profiling of neonates by mass spectrometry identified 882 metabolic features, and a set of 52 metabolites met the criteria of minimum similarity match and reverse match >600, S/N >10; metabolites contributed from column bleed were excluded. A close clustering of the quality control (QC) samples indicated minimal method-related variation, and outliers (CP: 1, CN: 2, NS: 2, HC: 1) were removed from further analysis (Figure 3A). The partial least squares-discriminant analysis (PLS-DA) plots clustered the CP and CN samples separately from the control (NS and HC) groups and between NS and HC (Figure 3B, 3C, 3D, Supplementary Figures S1A, S1B, S1C).

**Figure 3:**
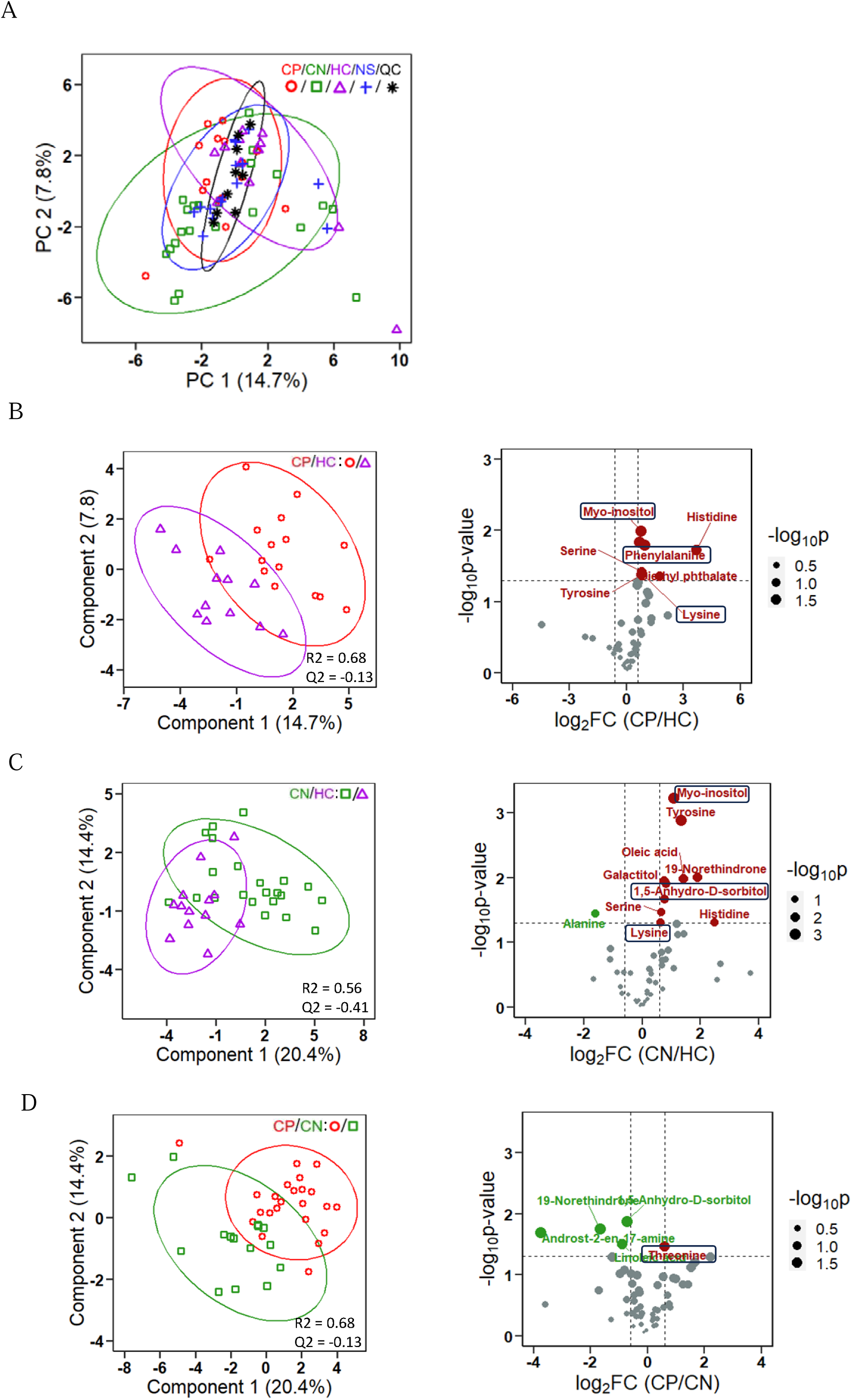
Serum metabolite profile of discovery set-I. **(A)** Principal Component Analysis (PCA) plot showing clustering of each group and QC samples. **(B-D)** Partial Least Square-Discriminant Analysis plot and volcano plot depicting group variance and significantly deregulated metabolic signatures (p<0.05, FC>1.5), respectively, between **(B)** CP and HC, **(C)** CN and HC, **(D)** CP and CN. CP: culture positive, n=16; CN: culture negative, n=24; NS: no sepsis, n=17; HC: healthy control, n=14; FC: Fold Change.

A seven deregulated (log2FC > 0.58, p < 0.05) metabolite signature, including phenylalanine, diethyl phthalate, histidine, myo-inositol, serine, tyrosine, and lysine, was identified as specific to CP compared to the HC group (Figure 3B). A metabolic signature of nine deregulated molecules (histidine, myo-inositol, serine, L-tyrosine, L-lysine, 19-norethindrone, oleic acid, galactitol, maltose) was identified as specific to the CN group (Figure 3C). A metabolite signature of five molecules (linoleic acid, 1,5-anhydro-D-sorbitol, 5α-androst-2-ene-17-one, threonine, 19-norethindrone) could differentiate CP from the CN sepsis groups (Figure 3D). The serum metabolic phenotype of the CP and NS groups was similar (Supplementary Figure S1A). However, 1,5-anhydro-D-sorbitol was deregulated in CN compared to the NS group (Supplementary Figure S1B). A metabolic signature of 6 metabolites (myo-inositol, oleic acid, talose, alanine, tyrosine, histidine) could differentiate NS from the HC group (Supplementary Figure S1C, S1D).

### Identification of serum metabolite biosignature that distinguishes neonatal sepsis from controls in discovery set-II

Gas chromatography-time of flight mass spectrometry (GC-TOF-MS) metabolite profiling of neonatal serum in discovery set-II identified 559 metabolic features, of which 57 met the set criteria for minimum similarity match and reverse match > 600, S/N > 10. Based on the QC sample clustering, the method-related variation was minimal (Figure 3A). A few samples from each study group (CP:1, CN:1; NS:1, HC:1) were identified as outliers and removed from further analysis. The PLS-DA model showed a distinct cluster of CP and CN sepsis groups compared to the control HC and NS groups and between NS and HC groups (Figures 4B, 4C, 4D, Supplementary Figures S2A, S2B, S2C).

**Figure 4:**
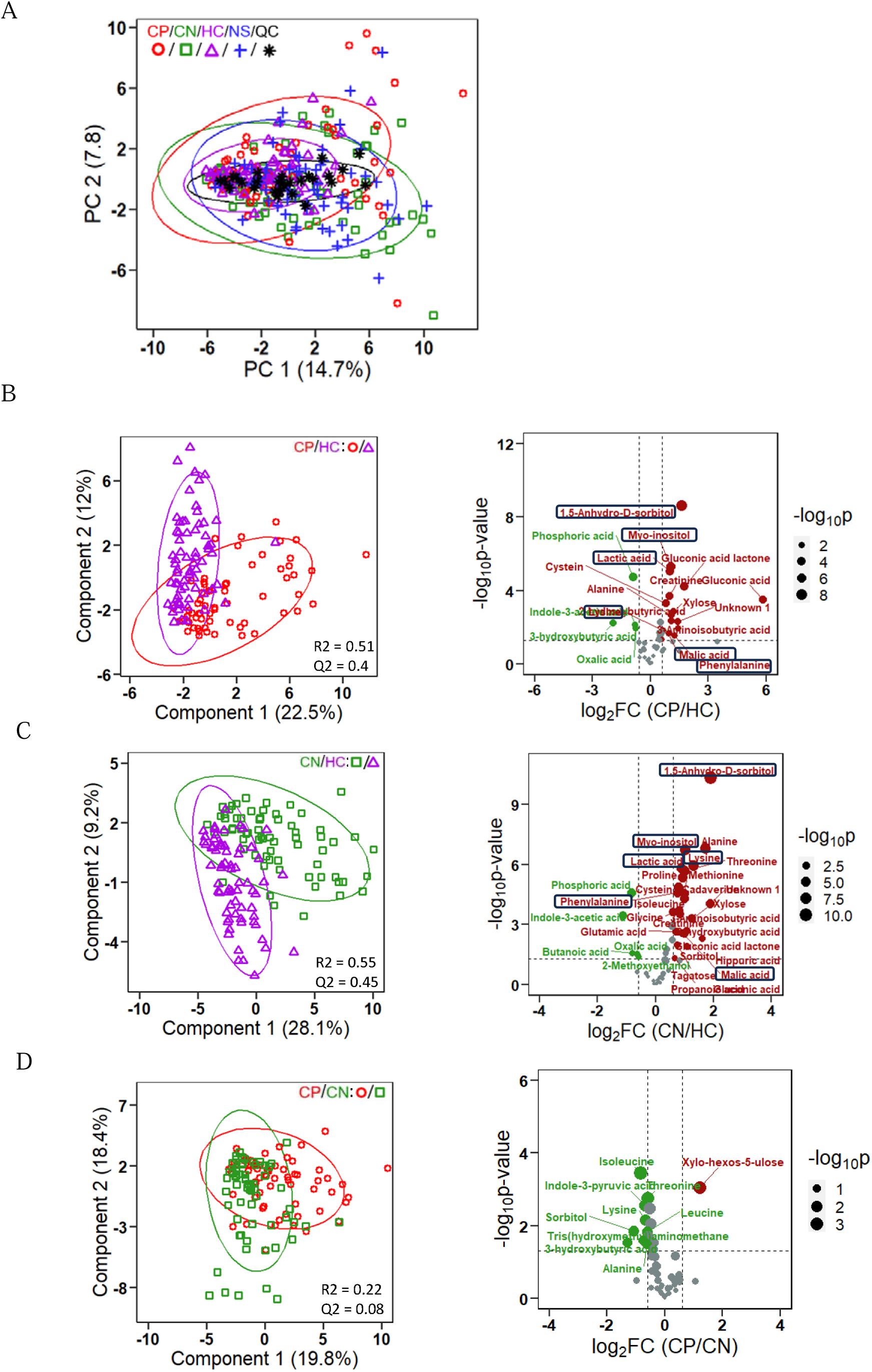
Serum metabolite profile of discovery set-II. **(A)** Principal Component Analysis (PCA) plot showing clustering of each group and QC samples. **(B-D)** Partial Least Square-Discriminant Analysis plot and volcano plot depicting group variance and significantly deregulated metabolic signatures (p<0.05, FC>1.5), respectively, between **(B)** CP and HC, **(C)** CN and HC, **(D)** CP and CN. CP: culture positive, n=63; CN: culture negative, n=69; NS: no sepsis, n=68; HC: healthy control, n=69; FC: Fold Change.

A set of 19 deregulated metabolites (1,5-anhydro-D-sorbitol, 2-hydroxybutyric acid, 3-aminoisobutyric acid, 3-hydroxybutyric acid, alanine, creatinine, cysteine, lactic acid, gluconic acid, gluconic acid-e-lactone, indole-3-acetic acid, lysine, malic acid, myo-inositol, oxalic acid, phenylalanine, phosphoric acid, xylose, Unknown 1) met the criteria and could distinguish the CP group from the HC group (Figure 4B). The serum metabolite profiles that distinguished *Klebsiella pneumoniae* and *Acinetobacter baumannii* infections from HC were similar, except for a few molecules (Supplementary Figure S3). Thirty-one deregulated metabolites were identified in CN compared to the HC groups (Figure 4C). Serum lysine, xylo-hexulose, and sorbitol showed deregulation in CP compared to NS (Supplementary Figure S2A). Eleven metabolites were deregulated between the CP and CN groups (Figure 4D, Supplementary Figure S2D, and Supplementary Table S5). Serum indole-3-acetic acid was deregulated in CN compared to NS (Supplementary Figure S2B). A metabolite signature of 25 metabolites could differentiate NS from the HC group (Supplementary Figure S2C).

A set of six metabolites (1,5-Anhydro-D-sorbitol, lactic acid, malic acid, myo-inositol, phenylalanine, and lysine) that could distinguish CP and CN from the HC group in discovery set I or II qualified as a sepsis biosignature and were selected for validation (Supplementary Table S6, Supplementary Figures S4A, S4B, S4C, S4D, S4E, S4F, S4G). The fragmentation patterns of the unknown metabolites 1 and 2 are shown in Supplementary Figure S4H and S4I. A perturbed inositol phosphate, ascorbate, and pyruvate metabolism was observed in the CP or CN sepsis groups compared to HC, based on the metabolite set enrichment analysis (MSEA) of both discovery sets (Supplementary Figure S5A, S5B). However, this six-metabolite serum biosignature showed the least correlation with white blood cell (WBC) count and total neutrophil count in neonates (Supplementary Figure S6A).

### Serum metabolite markers for predicting sepsis outcome

The serum glucose, gluconic acid, and cysteine levels were significantly higher (FC 2.0, Welch t-test, p-value <0.05) in the CP neonates who died from sepsis compared to those who survived (Supplementary Figure S6B). In the CN group, L-aspartic acid was lower in neonates who died than in the survivors (Supplementary Figure S6C).

### Performance evaluation of serum metabolite biosignature for sepsis diagnosis in an independent validation set

To quantify the predictive ability of the serum six-metabolite signature (1,5-anhydro-D-sorbitol, lactic acid, malic acid, myo-inositol, phenylalanine, and lysine), the area under the receiver operating characteristic curve (AUC of ROC/AUROC) was computed from the serum metabolites data of an independent validation set (n=60, CP/CN/NS/HC: 15 each). In CP, the AUROC was 0.97 against HC and 0.84 against NS (Figure 5A, Supplementary S7A). In CN, the AUROC was 0.97 against HC and 0.64 against NS (Figure 5B, Supplementary S7B). The metabolite signature, including sorbitol, lysine-3-hydroxybutyric acid, leucine, aminomethane, threonine, indole-3-pyruvic acid, isoleucine, xylo-5-hexosulose, and alanine, could distinguish CP from CN with an AUROC of 0.93 (Figure 5C). A separate model was built by adding confounding factors like gestational age and birth weight, but the predictive power of the molecular signature, based on the AUROC, was nearly the same across study groups (Supplementary Figure S8A, S8B, S8C, S8D).

**Figure 5:**
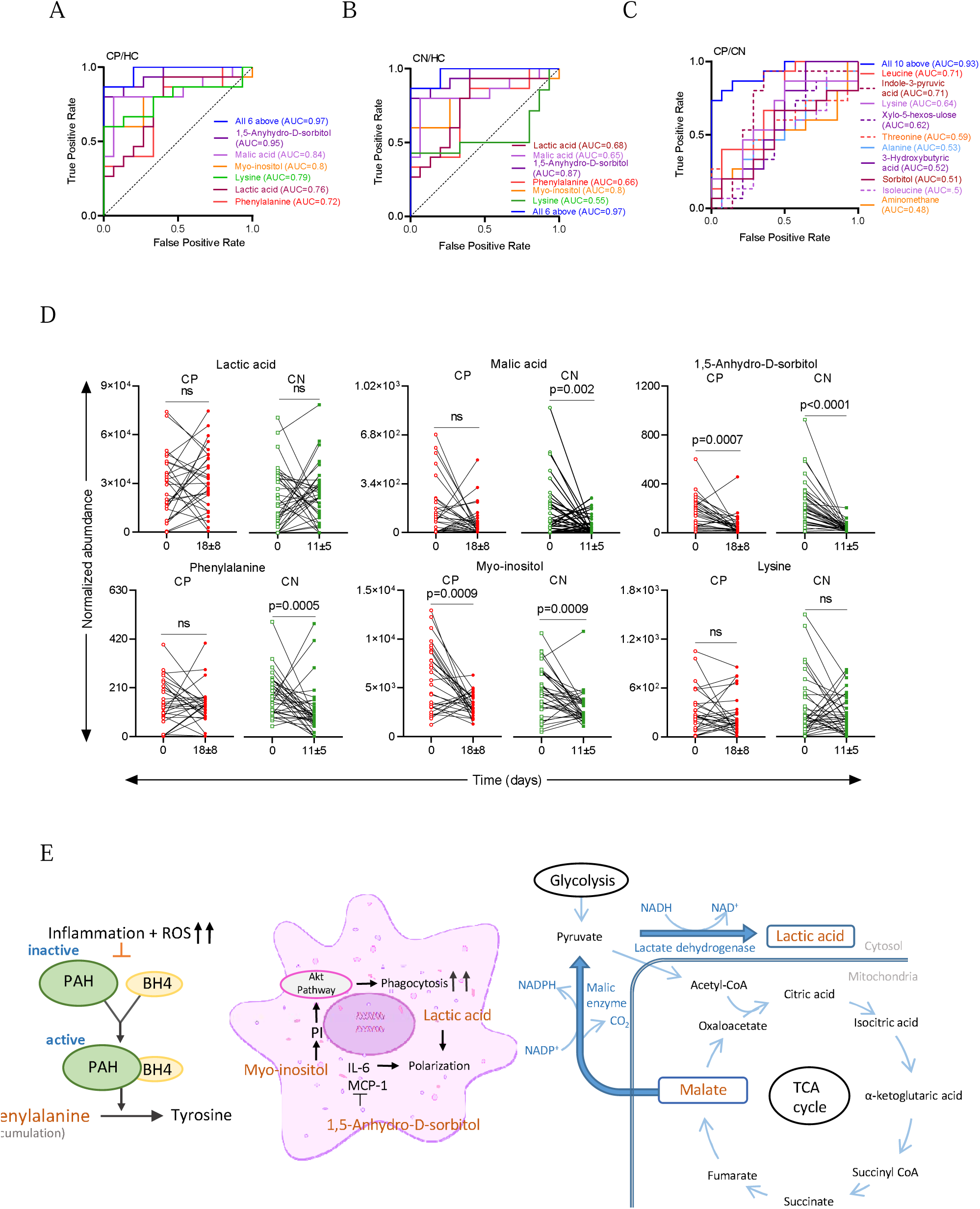
Validation of putative metabolic signature in an independent data set and their trend in followed-up sepsis cases upon recovery. Area Under the Receiver Operating Characteristic Curve (AUC of ROC) of the deregulated metabolite signature between (**A**) CP and HC group, (**B**) CN and HC group, (**C**) CP and CN group (CP/CN/HC:15 each). (**D**) Box plots depicting the abundance of individual deregulated metabolites in followed-up neonatal sepsis cases upon recovery (CP/CN:29/35). (**E**) The biological significance of the putative sepsis biomarkers. CP: Culture positive, CN: Culture negative, ns: not significant and p-value <0.05 is considered significant. PAH: Phenylalanine hydroxylase, BH4: Tetrahydrobiopterin, ROS: Reactive oxygen species, PI: Phosphatidyl-inositol, IL-6: Interleukin-6, MCP-1: Monocyte chemoattractant protein-1, NADH: Nicotinamide adenine dinucleotide, NADPH: Nicotinamide adenine dinucleotide phosphate.

### Targeted validation of the serum metabolite biosignature in follow-up cases of culture-positive, culture-negative, and no sepsis patients

In a separate follow-up group of sepsis cases and controls (n=100, CP/CN/NS: 29/35/36), the hospitalization duration was longer for CP and CN patients compared to NS (Figure 1B). Serum levels of 1,5-anhydro-D-sorbitol (p<0.0001) and myo-inositol (CP: p=0.002, CN: p=0.01), which were significantly higher at diagnosis, decreased to near-normal levels seen in healthy controls after recovery, that is, after completing antibiotic treatment (Figure 5D). Serum phenylalanine and malic acid levels normalized in the recovered CN and NS groups but did not significantly change in CP sepsis neonates. Serum malic acid showed a trend toward increase in recovered CP cases, although the difference was not statistically significant (Figures 5D, Supplementary Figure S9).

### Identification of serum markers to distinguish neonatal sepsis (CP/CN) from NS cases using LC-MS from an independent validation set-II

Since differentiating neonatal CP or CN groups from NS has greater clinical importance, a global serum metabolite profile was obtained using LC-MS on an independent validation set-II (n=100, including CP/CN/NS/HC, with 25 samples each) (Figure 6A). A total of 1615 metabolic features were identified, of which 151 met the set criteria (delta mass (ppm) >5 and <-5) and were selected for further analysis. Based on the clustering of QC samples, method-associated variation was minimal. The PLS-DA model showed clear clustering of CP and CN sepsis groups from the control NS group (Figure 6B). A five-metabolite signature, including PC(O-16:0/18:1(9Z)), PC(O-14:0/16:1(9Z)), PE(20:4(5Z,8Z,11Z,14Z)/0:0), 12-amino-dodecanoic acid, and 1,5-anhydro-D-sorbitol, can distinguish CP sepsis from the NS control group (Figure 6C). A set of three metabolites—PE(20:4(5Z,8Z,11Z,14Z)/0:0), 12-amino-dodecanoic acid, and 1,5-anhydro-D-sorbitol—can differentiate CN from the NS group (Figure 6C). A three-metabolite signature, consisting of PC(O-16:0/18:1(9Z)), PC(O-14:0/16:1(9Z)/0:0), and 12-amino-dodecanoic acid, can distinguish CP from the CN group (Figure 6D). A set of 5 serum metabolite signatures (PC(O-16:0/18:1(9Z)), PC(O-14:0/16:1(9Z)), hippuric acid, spermidine, and lactic acid) can distinguish the neonatal sepsis suspected groups (CP, CN, NS) from the HC controls (Figure 6D). Serum lactate levels can also differentiate the CP group from HC. Additionally, PC(O-16:0/18:1(9Z)), PC(O-14:0/16:1(9Z)), 12-amino-dodecanoic acid, hippuric acid, and spermidine can differentiate CN from HC. Similarly, PE(20:4(5Z,8Z,11Z,14Z)/0:0), hippuric acid, and 1,5-anhydro-D-sorbitol. differentiate the NS group from HC subjects (Figure 6D). The abundance of dysregulated serum metabolites identified in the LC-MS profiling of the validation cohort II samples was similar between neonates who succumbed to sepsis and those who survived (Supplementary Figure S10A). It also showed similar abundances between *Acinetobacter baumannii* and *Klebsiella pneumoniae* infections in the CP sepsis group (Supplementary Figure S10B).

**Figure 6:**
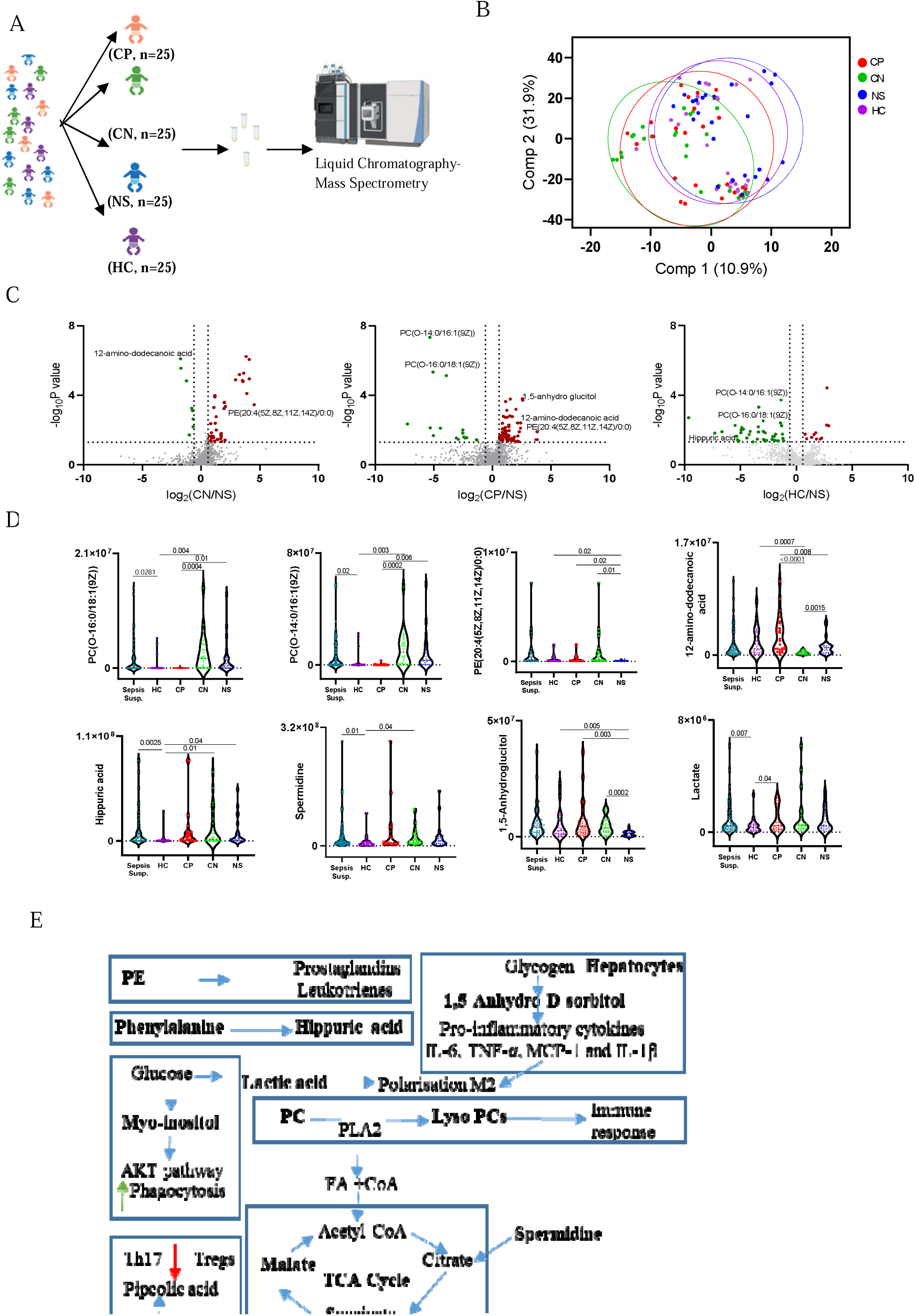
Serum metabolite profile using LC-MS of validation set-II. (**A**) Number of neonates included in validation set-II from each study groups. (**B**) PLS-DA plot depicting the clustering of study groups. (**C**) Volcano plots depicting the overall deregulated serum metabolites between study groups (CP/NS; CN/NS, HC/NS). (**D**) Violin plots highlighting key deregulated metabolites differentiating study groups with bars depicting the p-value between groups. CP: culture positive, CN: culture negative, NS: no sepsis, HC: healthy control. **(E)** Schematic representation of the metabolic pathways associated with the putative biomarkers identified by LC-MS analysis.

## DISCUSSION

Our metabolomic analysis successfully identified distinct metabolite profiles that can differentiate between clinical groups, with lactate serving as a key discriminator between the culture-positive (CP) sepsis and healthy control (HC) groups. Additionally, specific phospholipids (PC and PE species), amino acids, and organic acids, including hippuric acid, emerged as important biomarkers to distinguish between culture-negative (CN) and ‘no sepsis’ (NS) groups from HC, demonstrating the potential of metabolite-based markers for neonatal sepsis diagnosis or screening.

Neonatal sepsis can be caused by either mono-or polymicrobial infections, resulting in a complex immune response from the host.^12^ The pathogens or their molecular products disrupt host cell and tissue metabolism by affecting their biochemical processes. Such changes are reported to be reflected in the serum, urine, and cerebrospinal fluid (CSF) metabolic components as revealed by mass spectrometry analysis.^12-14^ We identified a serum biosignature consisting of 1,5-anhydro-D-sorbitol, lactic acid, lysine, malic acid, myo-inositol, and phenylalanine for neonatal sepsis cases. The lower AUC of ROC for the biosignature between CP/CN and NS may be due to the high similarity in their clinical manifestations. However, it still captured the difference between neonates with CP/CN sepsis and ‘no sepsis’. Factors like gestational age and birth weight did not significantly alter the predictive value of the existing models, suggesting that the molecular signature is independently associated with the disease. While our correlation analysis showed a significant relationship between age and the metabolite signature, this relationship does not appear to confound the association between the signature and the disease when directly included in the model.

Data acquisition for the two discovery sets was carried out using two instruments. As expected, gas chromatography coupled with time-of-flight mass spectrometry (GC-TOF-MS, used in discovery set-II) demonstrated a better detection range and higher sensitivity compared to GC coupled with quadrupole MS (GC-MS, discovery set-I). Because of this, data collection for validation set-I and follow-up samples was performed using GC-TOF-MS. Serum metabolite profiling using LC-MS offers the advantage of wider coverage and does not require chemical derivatization; which is necessary for GC-MS analysis and limits its coverage. LC-MS-based serum metabolite profiling in an independent validation set-II identified a set of metabolites (PC(O-16:0/18:1(9Z)), PC(O-14:0/16:1(9Z)), PE(20:4(5Z,8Z,11Z,14Z)/0:0), and 12-amino-dodecanoic acid) capable of differentiating CP from the NS group. Serum levels of 12-amino-dodecanoic acid, 1,5-Anhydro-D-sorbitol, and PE (20:4(5Z,8Z,11Z,14Z)/0:0) could distinguish neonatal CN cases from the NS group.

Phenylalanine metabolism is known to be upregulated in adult sepsis and is positively associated with a higher risk of mortality.^11,22,33^ Higher serum and plasma lysine levels are observed in adult patients and in the urine of neonates with sepsis.^24,25,26^ Lysine and phenylalanine are essential amino acids, and their uptake is vital for proper growth and development in neonates. An increased abundance of these amino acids could be linked to the reported intestinal dysbiosis seen in sepsis.^27^ Enhanced intestinal permeability due to gut dysbiosis increases the absorption of metabolites into the bloodstream. Impaired liver phenylalanine hydroxylase (PAH), which converts phenylalanine to tyrosine, may also lead to elevated serum phenylalanine levels in sepsis. This impairment could result from PAH’s dependence on its cofactor 5,6,7,8-tetrahydrobiopterin (BH4), which is susceptible to oxidative stress and often gets depleted during inflammatory conditions like sepsis (Figure 4E).^28^ Sepsis causes muscle atrophy, and lysine deficiency-induced apoptosis leads to protein breakdown in muscle cells, potentially raising serum lysine levels in neonates.^29,30^

High urine and serum myo-inositol levels were observed in neonatal and geriatric sepsis patients, respectively.^31^ Plasma myo-inositol levels are known to be positively correlated with the acute physiology score (APS) in patients with sepsis-induced acute lung injury (ALI).^32^ Myo-inositol helps depolarize the macrophage membrane potential, which enhances their phagocytic ability (Figure 5E).^33^ It is involved in the production of phosphatidylinositol (4,5)-bisphosphate (PIP2) and phosphatidylinositol (3,4,5)-trisphosphate (PIP3), which are associated with the *Akt* pathway. The *Akt* pathway activates macrophages and other immune cells and regulates apoptosis and oxidative stress in sepsis-induced tissue injuries.^34,35^

Serum 1,5-anhydro-D-sorbitol levels negatively correlate with blood glucose levels in diabetic patients.^36^ This is because 1,5-anhydro-D-sorbitol, a 1-deoxy form of glucose, is competitively inhibited by glucose during renal reabsorption.^37^ Uptake of 1,5-anhydro-D-sorbitol has been reported to reverse neonatal complications in infants of diabetic mothers.^38^ 1,5-anhydro-D-sorbitol is reported to block the production of pro-inflammatory cytokines such as IL-6 and monocyte chemoattractant protein-1 (MCP-1) in murine macrophages treated with lipopolysaccharide (LPS, Figure 5E).^39^ The higher levels of 1,5-anhydro-D-sorbitol in the CP and CN sepsis groups could result from anti-inflammatory responses by host immune cells against sepsis. Increased glycolysis during microbial infection may also contribute to high serum 1,5-anhydro-D-sorbitol levels due to increased cellular glucose uptake.^40^

Higher serum lactic acid levels are observed in sepsis patients and are directly linked to severity and mortality.^41^ Elevated lactic acid levels are a recognized marker for tissue hypoperfusion and hypoxia in sepsis, indicating metabolic disruption during the systemic inflammatory process.^42^ This occurs due to the Warburg effect in inflamed tissues and immune cells, where aerobic glycolysis produces adenosine triphosphate (ATP) to meet increased energy demands. The conversion of pyruvate to lactic acid is also increased to generate nicotinamide adenine dinucleotide (NAD), which supports glycolysis (Figure 5E). Lactic acid has been shown to promote M2-like macrophage polarization.^43^ High plasma malic acid levels have been found in patients with sepsis-associated encephalopathy.^44^ Besides its role as a tricarboxylic acid (TCA) cycle intermediate in mitochondria, malate is transported into the cytosol, where it converts to pyruvate, contributing to increased lactate production during inflammation.^41,45^ The malate-pyruvate shuttle also facilitates the production of nicotinamide adenine dinucleotide phosphate (NADPH), which is essential for glycolysis.^46^ Exposure to LPS has been reported to lead to the accumulation of malate, fumarate, and succinate in macrophages during inflammation.^47^

Serum hippuric acid can distinguish between neonatal sepsis groups (CP, CN, NS) and healthy controls in validation set-II, supporting earlier findings.^48^ Disrupted gut microbial metabolism or impaired liver clearance during infection contributes to increased hippuric acid levels.^48^ Phosphatidylcholines such as PC(O-14:0/16:1(9Z)) and PC(O-16:0/18:1(9Z)) are crucial for maintaining cell membrane integrity and signal transduction and were identified as markers for neonatal sepsis. Abnormal serum phospholipid levels suggest oxidative stress, immune cell activation, or endothelial dysfunction in sepsis.^49^ Serum 12-amino-dodecanoic acid, a medium-chain amino fatty acid, may reflect alterations in fatty acid metabolism or protein acylation during different stages of sepsis development and resolution, enabling differentiation among CP, CN, and NS groups. Serum PE (20:4(5Z,8Z,11Z,14Z)/0:0), a lysophosphatidylethanolamine of arachidonic acid, distinguishes CP and CN groups from NS, indicating differences in group-specific inflammatory mediator precursors and phospholipid remodeling.^50^ Derivatives of arachidonic acid are key to inflammatory mediators, and their disrupted metabolism in neonatal sepsis could influence immune activation states.^51^ Polyamines, especially spermidine, are elevated in neonates compared to adults and serve protective roles, including acting as antioxidants, reducing inflammation, inducing cytoprotective autophagy, and enhancing mitochondrial function.^52^

The elevated levels of phenylalanine, myo-inositol, malic acid, and 1,5-anhydro-D-sorbitol in CP, CN groups, and NS neonates decreased after treatment completion and recovery. The longer recovery time observed in the CP and CN groups could be due to the development of hyper-inflammation and subsequent immuno-paralysis, which are difficult to resolve.^53^ This validates their association with sepsis onset and suggests they could be targets for therapy development. Since most neonates with sepsis had EOS, the utility of the identified serum biosignature in late-onset sepsis requires further validation.

This study has several strengths and a few limitations. A key strength is the identification of serum metabolite markers from two discovery sets using mass spectrometry tools. The study involved 600 neonates from multiple clinical sites and adhered to strict diagnostic criteria to confirm culture-positive or culture-negative sepsis. Including a no-sepsis patient group with symptoms similar to sepsis as a control helps distinguish between neonates with suspected sepsis who are later found not to have sepsis and those with true sepsis. We employed appropriate statistical tools to identify the biomarker set, and the fact that these markers were also monitored in follow-up sepsis and no-sepsis subjects after treatment enhances the robustness of the findings. The origin of the identified metabolites could not be traced to the host, infectious agents, or gut microbial dysbiosis. However, the similarity between the serum metabolite profiles of CP cases with Klebsiella pneumoniae and Acinetobacter baumannii infections suggests that deregulated metabolites may result from the host’s inflammatory response.

Additionally, the correlation between markers like neutrophil count and WBC count, as well as other characteristics like birth weight and gestational age, and the serum metabolite signature was minimal (Supplementary Figure S6A). We could not correlate metabolite levels with disease severity because we did not record the Sequential Organ Failure Assessment (SOFA) score of the neonates. Although a few serum metabolites could distinguish surviving neonates from those who eventually died, their levels could not be measured in the serum of deceased neonates due to ethical concerns. Identifying key enzymes that catalyze the deregulated pathways contributing to the variations in the serum metabolic signature will be essential for better understanding disease mechanisms and developing therapeutics.

To conclude, this study identified a serum biosignature comprising myo-inositol, phenylalanine, malic acid, lysine, lactic acid, and 1,5-anhydro-D-sorbitol, which can accurately differentiate neonates with sepsis from healthy controls. A metabolic signature including PE(20:4(5Z,8Z,11Z,14Z)/0:0), 12-amino-dodecanoic acid, and 1,5-anhydro-D-sorbitol could distinguish culture-positive and culture-negative sepsis from the ‘no sepsis’ group. Much of this conclusion may be attributed to gut dysbiosis and underscores the importance of the gut microbiome in sepsis. The identified disrupted metabolic pathways offer critical targets for developing host-directed therapeutics in cases of neonatal sepsis, where limited options exist. Further research is necessary to uncover the molecular mechanisms behind these altered sepsis-specific serum metabotypes and to explore the development of therapeutics tailored for neonatal sepsis to enhance their treatment.

## MATERIALS AND METHODS

### Subject recruitment and classification

The study received approval from the Institutional Ethics Committees of the All India Institute of Medical Sciences (IEC-1074/06.11.20, RP-28/2020), Vardhman Mahavir Medical College (VMMC) and Safdarjung Hospital, New Delhi (IEC/VMMC/SJH/Project/2021-05/CC-157, dated 24.07.2021), Dr. Baba Saheb Ambedkar Medical College and Hospital (5(32)/2020/BSAH/DNB/PF, dated 31.08.2020), Lady Hardinge Medical College (LHMC/IEC/2022/03/30), the University College of Medical Sciences & Guru Teg Bahadur Hospital (IECHC-2022-51-R1, dated 07.12.2022), and the International Centre for Genetic Engineering and Biotechnology (ICGEB), New Delhi (ICGEB/IEC/2021/28, version 2). Preterm neonates born between 28 and 34 weeks of gestation and admitted to the neonatal intensive care unit (NICU) at the clinical sites from July 2022 to December 2023 were enrolled after obtaining written consent from parents or legal guardians. Neonates born to human immunodeficiency virus (HIV) +ve or hepatitis +ve mothers, or those with a postnatal age of over 28 days, were excluded (Figure 1A, Supplementary Table S7). Neonates born without complications or sepsis symptoms within 5 days of birth were recruited as HC from each clinical site.

The clinical team, along with the research staff, monitored all enrolled neonates for sepsis. Sepsis was suspected when there were either maternal or perinatal risk factors or clinical signs (Supplementary Table S7). The research nurses performed the diagnostic work-up for sepsis, including a sepsis screen, at the time of suspicion and before starting antibiotics. Whole blood sample (1 ml) was collected from the neonates for culture in automated culture bottles (BD BACTEC or BACT/ALERT) and incubated at 37□ before being transported to the microbiology laboratory within 12 to 24 hours of collection.

Based on the laboratory test results and the Centers for Disease Control and Prevention’s (CDC) National Healthcare Safety Network (NHSN) infection definitions, the neonates were categorized as having CP sepsis, CN sepsis, NS, or HC (Supplementary Table S8).^54^ Primarily, neonates with a positive screen whose blood culture grew a true pathogen were classified as CP, while those with sterile cultures or cultures growing commensals were grouped as CN. The sepsis screen included: white blood cell count < 4.0 × 10^9^ cells/L; absolute neutrophil count < 1.5 × 10^9^ cells/L; I/T ratio > 0.2; CRP > 6 mg/L; TLC ESR > 15 mm in the first hour; and the screen was considered positive if at least two of these findings were abnormal.^55^ Neonates suspected of sepsis but not meeting the criteria for CP or CN sepsis were classified as NS (Supplementary Table S8). The decision to initiate antibiotics was made at the discretion of the clinical team once the samples for the sepsis workup were collected, in accordance with their unit’s policy.

Random sampling using an online randomizer (www.randomizer.org) was performed to assign the enrolled neonates into the discovery set-I (n=71, 59.2% female, CP/CN/NS/HC: 16/24/17/14), discovery set-II (n=269, 43.4% female, CP/CN/NS/HC: 63/69/68/69), and validation set-I (n=60, ∼40% female, CP/CN/NS/HC: 15 each), as well as validation set-II (n=100, 42% female, CP/CN/NS/HC: 25 each). An independent set of neonates (n=100, CP/CN/NS: 29/35/36) not used in the earlier discovery set-I and -II was followed up until recovery to monitor the levels of deregulated metabolites.

### Serum Sample Processing

A part of the whole blood (0.5 ml) was transferred to serum collection tubes and kept still for 10-20 minutes. After centrifuging at 1,000 × g for 10 minutes at 4□, the supernatant was transferred into fresh tubes before storing at -80□. Each set’s quality control (QC) sample was prepared separately by pooling equal volumes of serum samples. All samples from the participating subjects were randomized using the online tool and run in batches of 20 with at least 2 QC samples per batch.

### Serum sample processing and metabolite extraction

A set of randomized stored serum samples (n=20) with 2 QCs were thawed at 4□, and to an aliquot (40 μl) of it, chilled methanol (80%, 320 μl) was added with a spike in standard (ribitol: 0.5 μg/μl, 1 μl). After incubating it in ice for 30 minutes, it was centrifuged at 4□ for 10 minutes at 15,000 × g. The supernatant was transferred to a fresh methanol-treated centrifuge tube. A blank tube was used as an extraction blank with ribitol spiked in. After drying the samples in a Speedvac (Labconco, USA), they were derivatized using N-Methyl-N-(trimethylsilyl) trifluoroacetamide (MSTFA). After adding (R)-2-Methoxy-N-(piperidin-3-yl)acetamide hydrochloride (MOX-HCl) solution (20 mg/mL; 40 μl) to each sample tube, they were incubated at 60□ for 2 hours at 900 rpm. Then MSTFA (70 μl) was added to each sample tube and incubated at 60□ for 30 minutes at 900 rpm. The derivatized samples were centrifuged at 16,000 × g for 10 minutes at room temperature, and the supernatant (∼90 μl) was transferred to fresh GC vials with 250 μl glass inserts for analysis using different GC-MS platforms.

### GC-MS Data acquisition

The discovery set-I samples were analyzed using a GC coupled to a quadrupole-MS (the Agilent 5975c TAD series). Using helium gas at a constant flow rate of 1 ml/minute, the derivatized sample (1 μl) was injected, in splitless mode, into an RTX-5 column (30 m by 0.25 mm by 0.25 μm; Restek, USA). The oven temperature gradient was set from 50□ to 280□ with a ramp of 8.5□/minute from 50 to 200□ and 6□/minute from 200 to 280□ with a hold of 5 minutes for metabolite separation. The temperature of the MSD transfer line was set at 260□ with a holding time of 1.37 minutes. The detector voltage was set at 1388 V, and a mass range of 50 to 600 m/z was selected. The MS ion source and quadrupole temperatures were set at 230□ and 150□, respectively. A solvent delay of 10 minutes was set, and the total GC-MS run time was 36.98 minutes.

For discovery set-II, validation, and follow-up sets, the derivatized test samples were analyzed using a GC-TOF-MS instrument (Pegasus 4D; Leco, USA). Using helium gas at a constant flow rate of 1 ml/minute, the derivatized sample (1 μl) was injected, in splitless mode, into an RTX-5 column (30 m by 0.25 mm by 0.25 μm; Restek, USA). The oven temperature gradient was set from 50□ to 280□ with a ramp of 8.5□/minute from 50 to 200□ and 6□/minute from 200 to 280□ with a hold time of 5 minutes for metabolite separation. The detector voltage was set at 1400 V, and a mass range of 30 to 600 m/z was used for detection. The temperature of the MSD transfer line was set at 250□, and the flow rate was set at 1 ml/minute. The MS ion source and acquisition rate were set at 220□ and 30 spectra/second, respectively. The total GC-MS run time was 36.98 minutes with a 10 minutes solvent delay.

### GC-MS Data Pre-processing

The .d files from the discovery set-I were then imported to the ChromaTOF^®^ (LECO, USA) software for further analysis. Using the “Statistical Compare” feature, the raw data files obtained from each data set were separately analysed in the ChromaTOF® (LECO, USA) software. Data matrices consisting of the identified metabolites as variables and the peak areas identified in the samples of each dataset were created. The peak areas were normalized using the internal standard i.e. ribitol. NIST 11 (National Institute of Standards and Technology, 243893 spectra; 30932 replicates) Mass Spectral database and LECO/Fiehn Metabolomics Library (ChromaTOF, USA) were used for the tentative identification of the molecular features. The minimum similarity match was set to 600 for annotation. The identification of significantly deregulated features was further confirmed by running commercial standards using the same GC-MS parameters.

### Serum metabolite extraction for LC-MS analysis

Serum samples (50 µL) of a subset of neonatal sepsis cases and controls were processed by adding chilled methanol (80 %, 200 µL), vortexed for 10 seconds, and incubated at –20°C for 10–12 hours for metabolite extraction. These processed samples were centrifuged at 16,000 × g for 10 minutes, and the supernatant was transferred to a fresh tube for drying using a Speed Vac. The dried samples were reconstituted in acetonitrile (105 µL, 5%) with an internal and external standard. An equal amount of extracted metabolites from each sample was pooled to prepare a quality control (QC) sample.

### LC-MS data acquisition

The extracted serum metabolites from validation set-II (n=109, samples/QC:100/9) were run in batches of 20 randomized samples, and a separate QC was run before and after completing data acquisition. Metabolomic analysis is performed on the Thermo Scientific^TM^ UHPLC system combined with Q Exactive Orbitrap Mass Spectrometer in both positive and negative ion modes. A C18 column, i.e., Hypersil GOLD (100 x 2.1 mm, 3.0 µm particle size, Thermo Fisher Scientific, USA), was used for separating the metabolites with a gradient of Milli-Q water (solvent A) and acetonitrile (solvent B) as eluents, containing 0.1% formic acid, and the total run time was 25 minutes. The gradient begins with 5% of solvent B, then increases to 99% at 17 mins, is maintained at 99% till 21 mins and is reduced to 5% at 22 mins and is maintained till the end of the run with a flow rate of 0.5 mL/min. The resolution of the mass spectrometer was set at 70,000 for full MS and 17500 for ddMS2 and scanned at a mass range of 150 to 2000 m/z. The capillary temperature was 340□, Sheath Gas Flow rate was set at 54, Aux Gas rate was set at 13, and ion spray voltage was set at 5.5 kV for positive mode and 4.5kV for negative mode. The ion source temperature was set at 250/300□ (Positive/Negative).

### LC-MS data analysis

The raw LC-MS data files obtained from QExactive were analyzed using Compound Discoverer software (version 3.3 SP3, Thermo Fisher) for chromatography peak alignment, mass spectrum visualization, metabolite identification, and statistical analysis. Peaks with a mass tolerance of 5 ppm, signal noise ratio (S/N) of 3, and a minimum peak intensity of 10^6^ were identified using The ChemSpider, comprising BioCyc, KEGG, and Human Metabolome Database (HMDB). To assign compound annotation on the MS/MS level, mzCloud, ChemSpider, and Metabolika searches with a mass tolerance of 5 ppm were used. Out of the total 1615 metabolites, metabolites with delta mass (ppm) greater than 5 and less than -5 are removed. The differentially regulated metabolite IDs were extracted from ChemSpider, HMDB, KEGG, CHeBI, CAS, and Drugbank databases. For pathway analysis, websites like Impala, Reactome, and MetScape are used.

### Multivariate and univariate statistical analysis

Clinical parameters with p<0.05 at a 95% confidence interval between study groups were selected as significant using the Wilcoxon ranked-sum test. MetaboAnalyst 4.0, GraphPad Prism 9.0.0 (GraphPad Software, Boston, Massachusetts, USA), and RStudio were used to analyze meta data generated from the mass spectrometry to identify deregulated metabolites.^56^ PCA and PLS-DA model were generated separately for each dataset using MetaboAnalyst. A Welch’s t-test (unequal variance of the two groups) was performed to compare the differences in the abundance of all the metabolites between the groups using normalized data. A paired *t*-test was performed for the follow-up data to monitor the abundance of the deregulated metabolites. All identified metabolites were selected for MSEA using MetaboAnalyst to identify the perturbed molecular pathways. The AUC of ROC was calculated using the “combiroc” package, and Pearson’s correlation analysis was performed using “corrplot” in R. For visualization of the resulting data, different software – GraphPad Prism 9, Microsoft PowerPoint, and R packages “ggplot2” and “scatter3d” – were employed. An overview of the workflow is represented in Figure 1B. Metabolites with a fold change value >±1.5 and Welch’s t-test p-value <0.05 were considered significantly deregulated metabolites. For validation set-II, the molecules qualifying a VIP score>2.0 from the PLS-DA model, log_2_FC>±0.58, and Welch’s t-test p-value <0.05 were selected as differentially expressed metabolites.

## DATA AVAILABILITY

All datasets used for analysis are at DOI: 10.17632/3xwzkwf9g7.1 (https://data.mendeley.com/datasets/3xwzkwf9g7/1).

## ACKNOWLEDGEMENTS

We thank the hospital staff of the clinical sites for helping us with sample collection and preprocessing. We would like to acknowledge Dr Sushil Shrivastava, Dr Meetu Salhan, Dr Harsh Chellani, Dr Pratima Anand, and Dr Narendra Pal Singh for their constant support and involvement in this project, and Vrinda S Nair for assistance with the manuscript submission.

We would also like to thank the Department of Biotechnology (DBT), India, for financial support. The core support from the International Centre for Genetic Engineering and Biotechnology, New Delhi, to the Translational Health Group to maintain the Gas Chromatography and Mass Spectrometry facilities is acknowledged. We also thank the Translational Health Group members for their help and support.

## AUTHOR CONTRIBUTIONS

Conceptualization: R.N. Experiment design: R.N., R.A. Methodology: A.S., R.A., S.T., N.Y., V.S.N. Provided clinical samples and data: P.D., R.G., G.P.K., R.G., R.S., K.N., R.K., S.N., V.K. Sample storage and record keeping: R.A., A.S., S.T. Clinical data management and analysis: R.A., A.B. Sample processing: R.A., S.T., V.S.N. Method standardization: R.A., A.S. Data acquisition: R.A., A.B., S.T., N.S., J.S.M. Data analysis: R.A., S.T., N.Y., S.K.M., S.N. Investigation: R.A. Supervision: R.N., M.J.S. Funding acquisition: R.N., R.G., R.K., S.N., G.P.K., R.G., R.S., K.N., K.A., M.J.S. Coordination and strategy: R.N., M.J.S. Writing-original draft: R.A., R.N. Writing-review and editing: R.A., R.N., S.T., N.Y., V.S.N., R.G., R.K., S.N., G.P.K., R.G., R.S., K.N., M.J.S., S.K.M., K.A. All authors read and approved the final manuscript and had full access to all the data in the study.

## MATERIALS AND CORRESPONDENCE

RN and MJS

## SUPPLEMENTAL INFORMATION

**Supplementary File.** Supplementary Figures S1–S10, and Supplementary Tables S2–S8: Supplementary Figure S1. Serum metabolite profile of CP, CN, NS neonatal sepsis cases and HC in discovery set-I.

Supplementary Figure S2. Serum metabolite profile of CP, CN, NS sepsis cases and HC in discovery set-II.

Supplementary Figure S3. Metabolite profile of CP neonates infected with *Acinetobacter baumannii* and *Klebsiella pneumonia* against HC show pathogen-specific differences.

Supplementary Figure S4. Confirmation of the identity of the key deregulated metabolites in neonatal sepsis subjects by running commercial standards and fragmentation patterns of the deregulated unknown analytes.

Supplementary Figure S5. Metabolite Set Enrichment Analysis (MSEA) between sepsis and control groups.

Supplementary Figure S6. Correlation between sepsis serum metabolite signature and putative serum metabolite markers for disease severity.

Supplementary Figure S7. AUC of ROC of the biosignature consisting of 6 biomarkers depicts high predictive ability for CP and CN cases against the NS group.

Supplementary Figure S8. AUC of ROC of serum metabolic biosignature considering their birth weight and gestational age.

Supplementary Figure S9. The trend in the abundance of deregulated metabolites in longitudinally followed-up NS cases upon recovery.

Supplementary Figure S10. Deregulated serum metabolites and their distribution in culture-positive neonatal sepsis subjects in validation set-II.

Supplementary Table S2. Group-specific epidemiological details and their significance in neonatal sepsis and control cases.

Supplementary Table S3. Distribution of *Acinetobacter baumannii* and *Klebsiella pneumoniae* cases in the culture-positive neonatal sepsis cases.

Supplementary Table S4. Study group specific distribution of gestational age and birth weight of neonatal sepsis cases (CP/CN: culture +/-ve) and controls (NS/HC: no-sepsis and healthy controls).

Supplementary Table S5. Fold change (FC) and (p-value) of deregulated serum metabolites that could differentiate sepsis from control groups.

Supplementary Table S6. List of the serum metabolites selected for biomarker identification and validation

Supplementary Table S7. The criteria used for neonatal sepsis suspicion, confirmation and grouping are adopted in this study.

Supplementary Table S8. Definition used for classification of the neonatal sepsis suspected cases.

**Supplementary Table S1.** The Table containing individual patient details is presented in a separate Excel file.

