## Supplemental Material for "Discovery and validation of serum metabolic signature of neonatal sepsis"

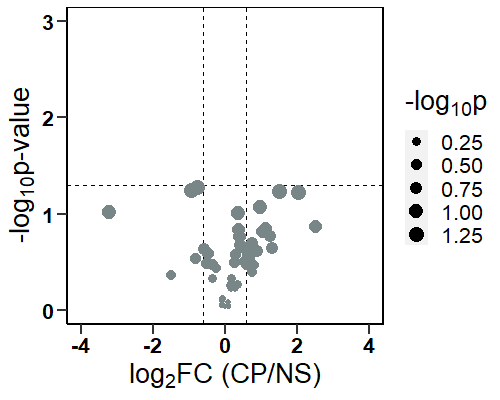

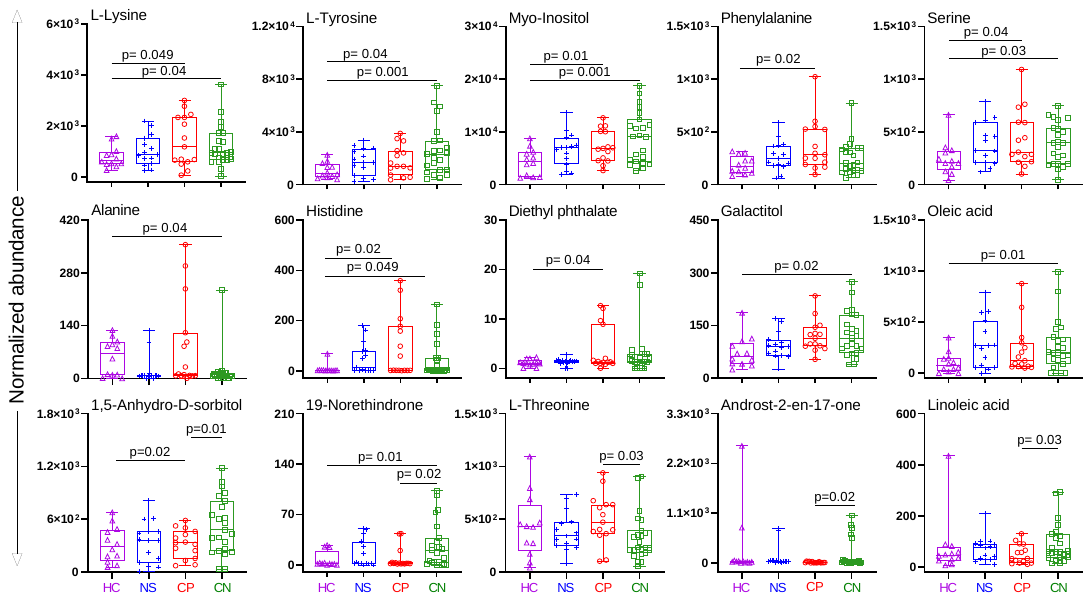

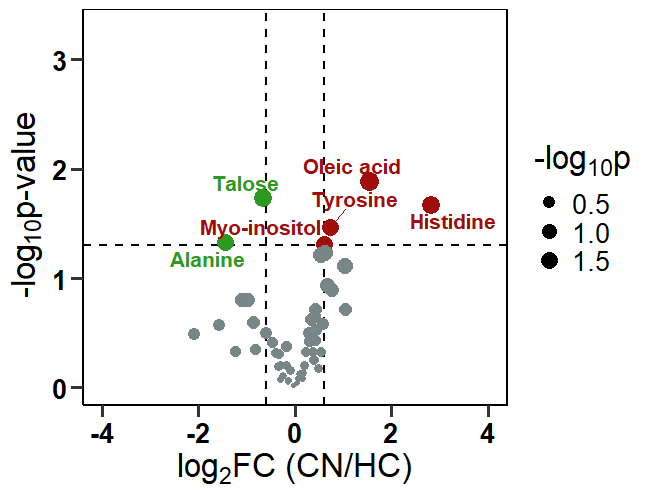

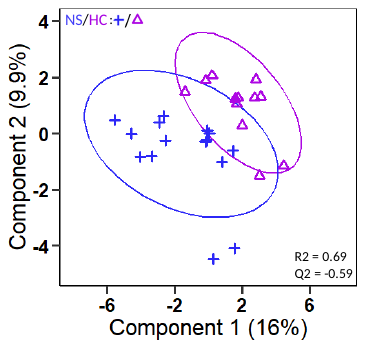

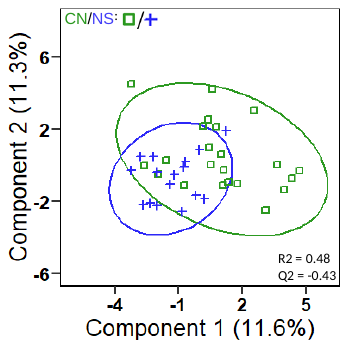

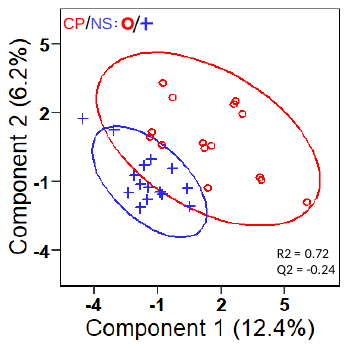
**Fig. S1.**

D

C

B

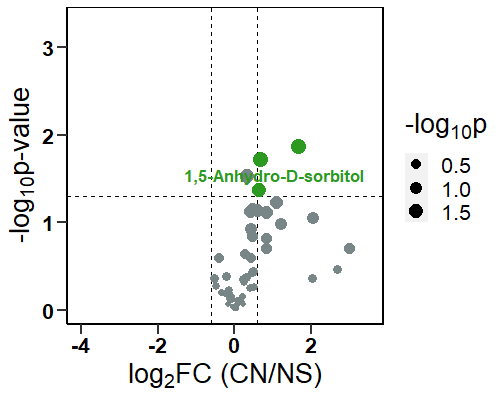

A

**Supplementary Figure S1. Serum metabolite profile of CP (n=16), CN (n=24), NS (n=17) neonatal sepsis cases and HC (n=14) in discovery set-I.** (**A-D**) Partial Least Square-Discriminant Analysis (PLS-DA) model and box plot depicting the group-specific variation and deregulated metabolites between CP and NS **(A)**, CN and NS **(B)**, and NS and HC **(C)**. (**D**) Box plots display the abundance of deregulated metabolites among CP, CN, NS and HC groups. CP: culture-positive, CN: culture-negative, NS: no sepsis, HC: healthy control.

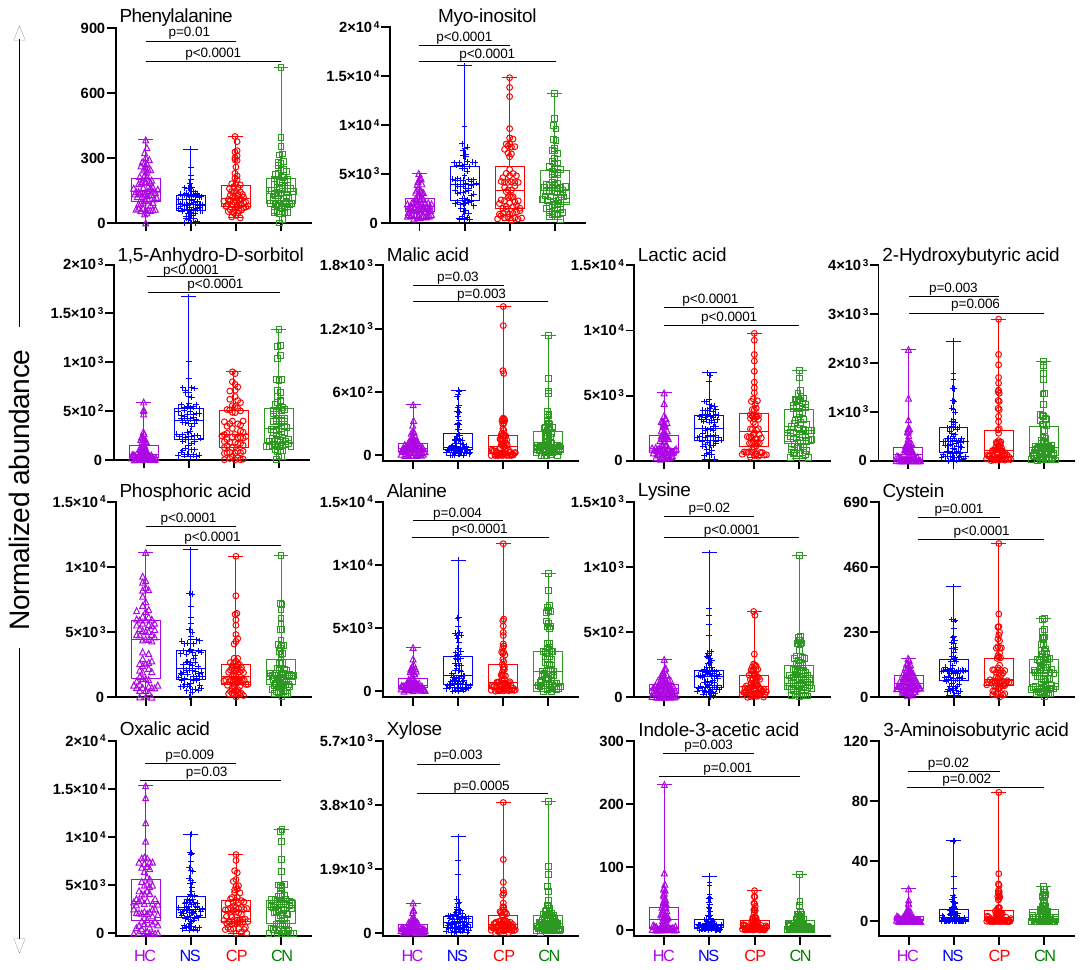

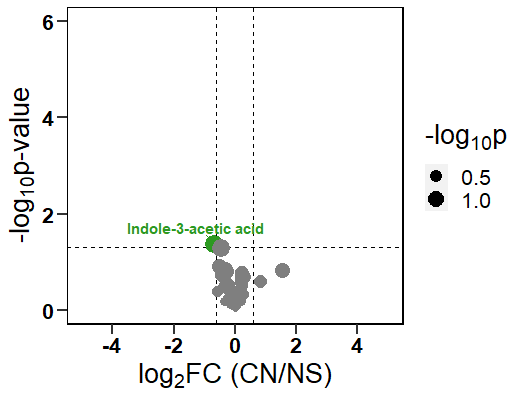

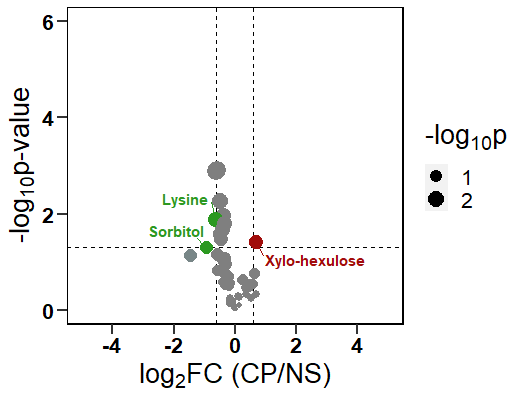

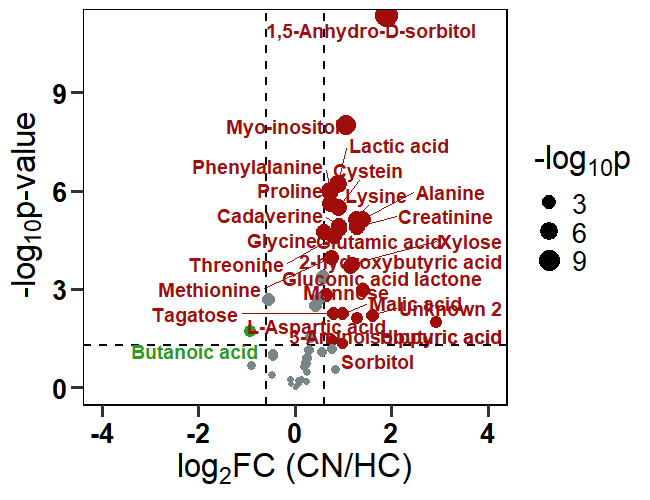

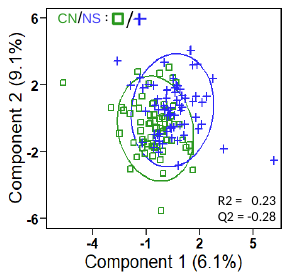

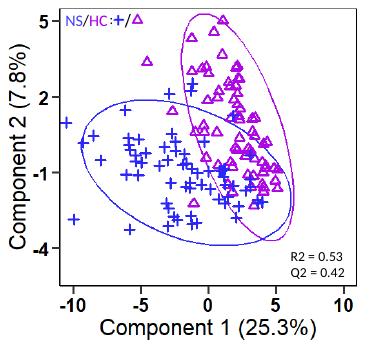

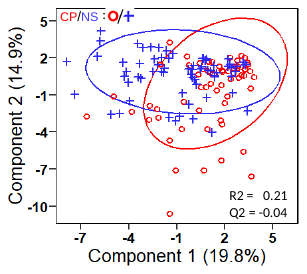
**Fig. S2.**

D

C

B

A

**Supplementary Figure S2. Serum metabolite profile of CP (n=63), CN (n=69), NS (n=68) sepsis cases and HC (n=69) in discovery set-II. (A-D)** Partial Least Square-Discriminant Analysis (PLS-DA) model and box plot depicting the group-specific variation and deregulated metabolites between CP and NS **(A)**, CN and NS **(B)**, and NS and HC **(C)**. (**D**) Box plots display the abundance of deregulated metabolites among CP, CN, NS and HC groups. CP: culture-positive, CN: culture-negative, NS: no sepsis, HC: healthy control.

**Fig. S3.**

Supplementary Figure S3. Metabolite profile of CP neonates infected with *Acinetobacter baumannii* and *Klebsiella pneumonia* against HC show pathogen-specific differences. Volcano plots show differences in the metabolite profile between *Acinetobacter baumannii* and HC (A) and *Klebsiella pneumoniae* and HC (B). Ab: *Acinetobacter baumannii;* Kp: *Klebsiella pneumonia;* HC: healthy control.

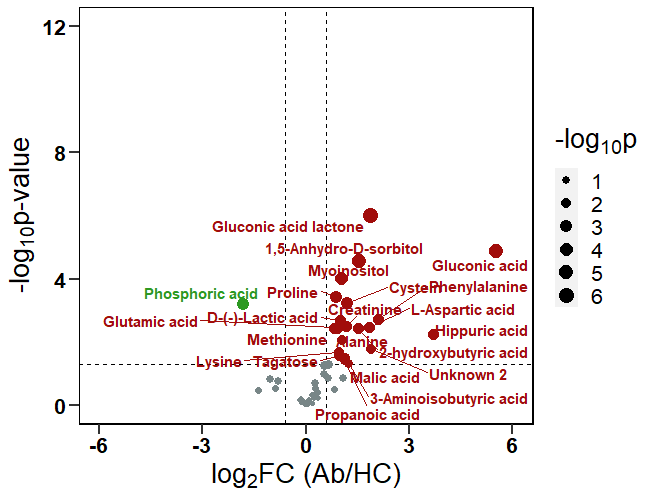

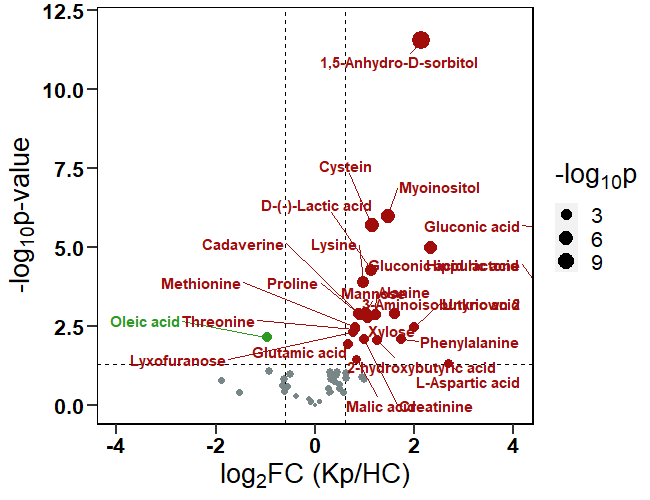
Fig. S4.

A

B

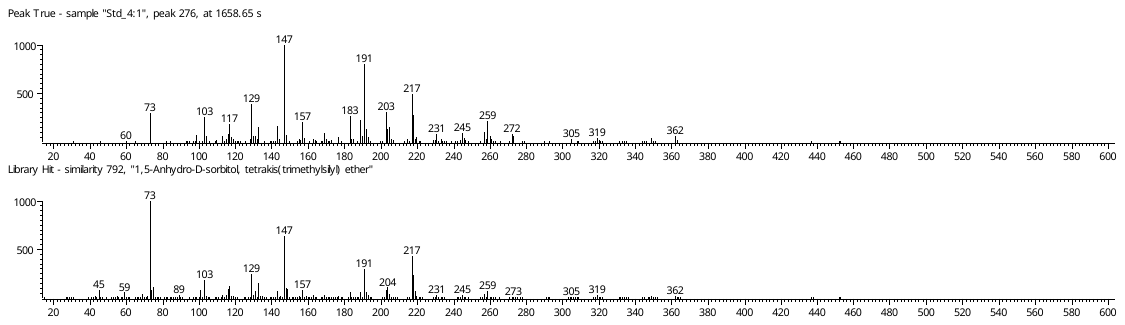

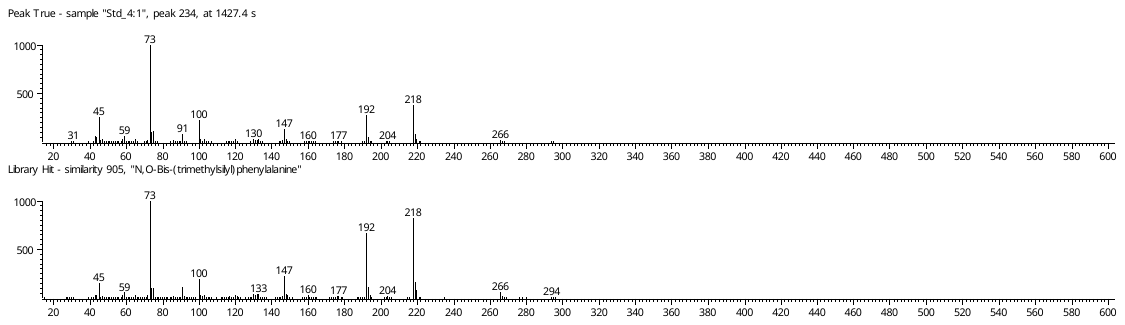

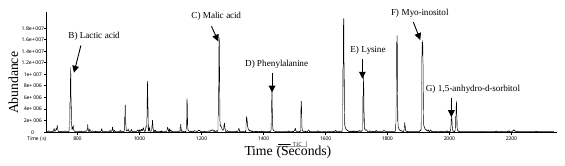

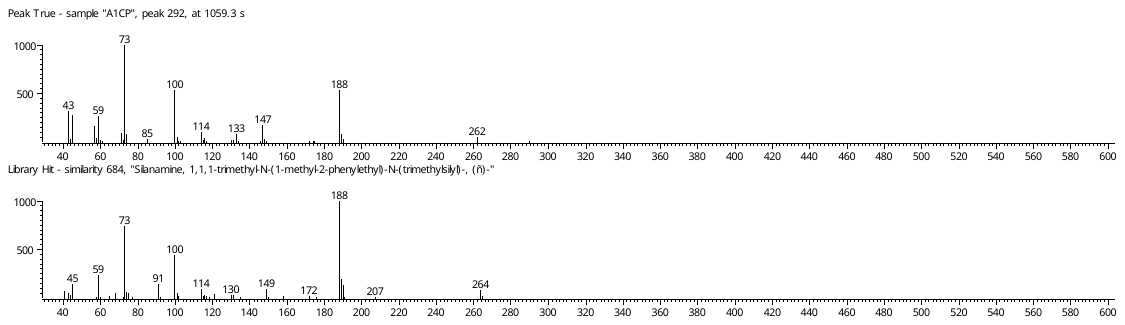

A

C

D

E

F

G

B

H

I

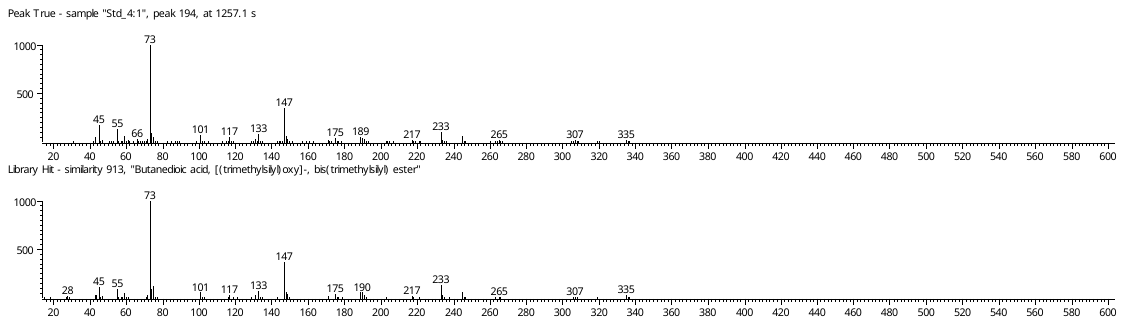

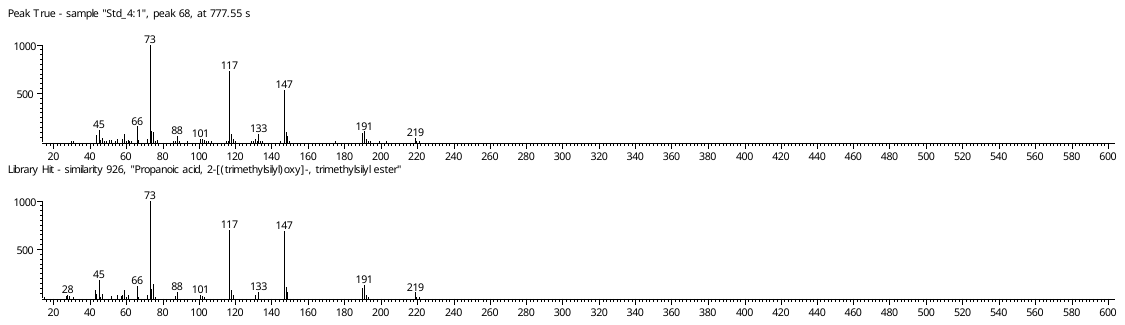

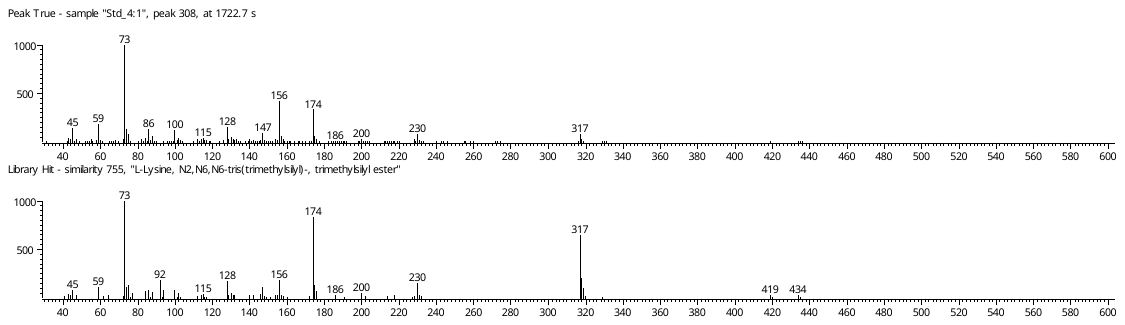

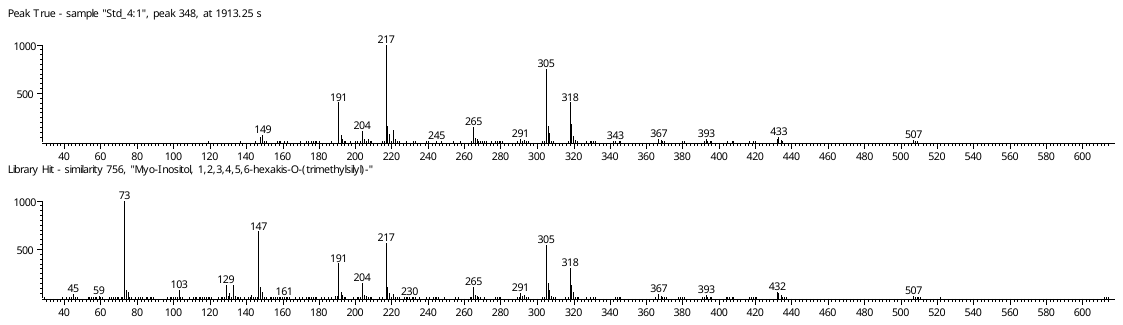

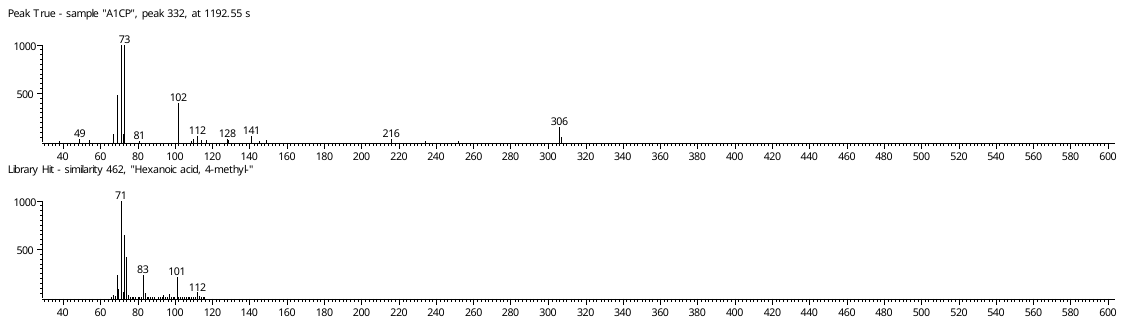

**Supplementary Figure S4. Confirmation of the identity of the key deregulated metabolites in neonatal sepsis subjects by running commercial standards and fragmentation patterns of the deregulated unknown analytes. (A)** Total ion chromatogram (TIC) of standard molecular mix spiked into serum samples reveals the identity of deregulated metabolites. **(B-G)** Fragmentation pattern of **(B)** lactic acid, **(C)** malic acid, **(D)** phenylalanine, **(E)** lysine, **(F)** myo-inositol, **(G)** 1,5-Anhydro-D-sorbitol trimethylsilyl (TMS) derivatives, and the details of **(H)** Unknown 1, and **(I)** Unknown 2 from the samples.

Fig. S5.

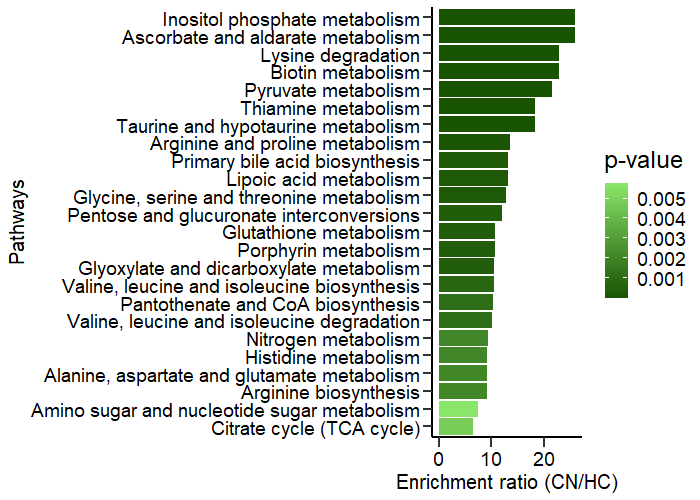

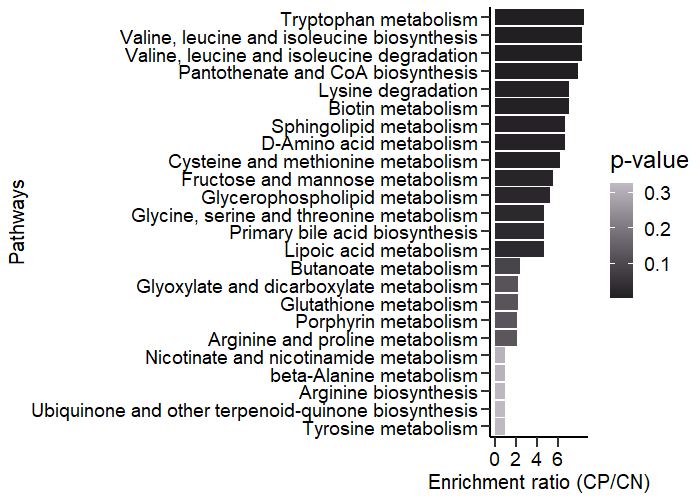

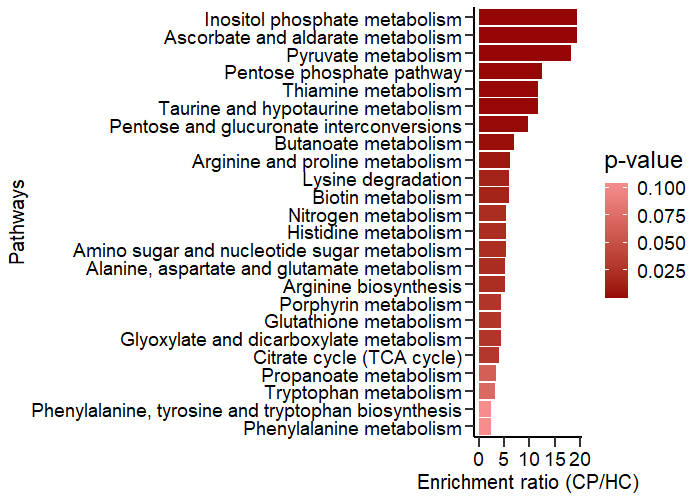

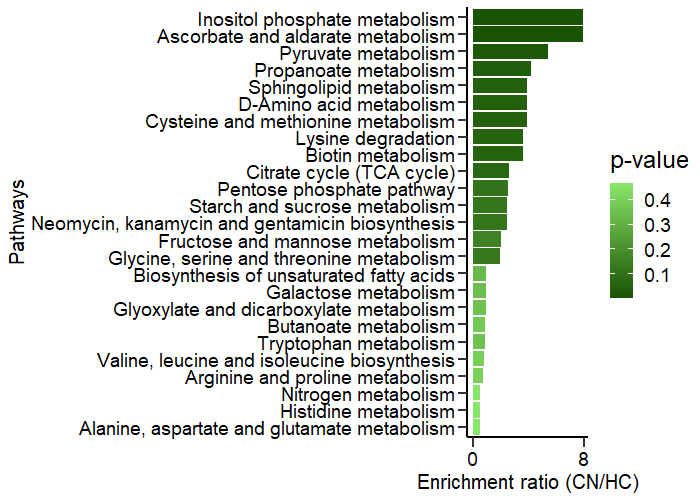

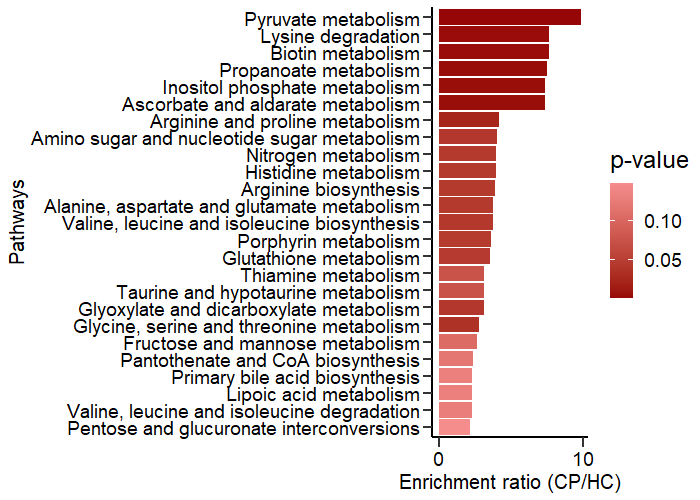

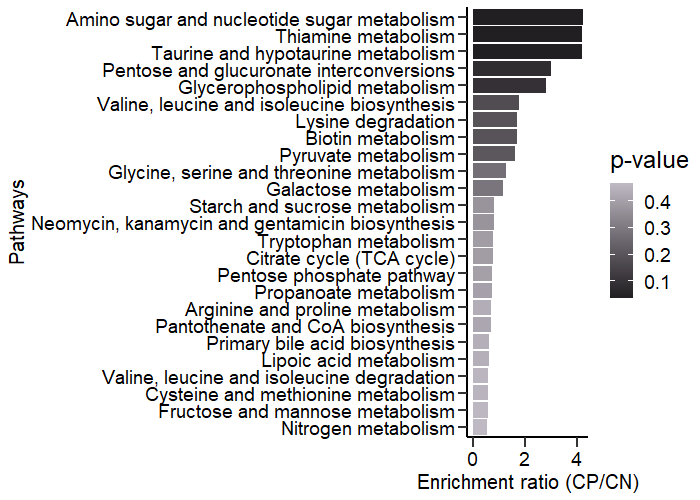

B

A

**Supplementary Figure S5. Metabolite Set Enrichment Analysis (MSEA) between sepsis and control groups.** MSEA plot depicting the deregulated pathways between sepsis (CP, CN) and control groups (HC) in Discovery set-I **(A)**, and Discovery set-II **(B)**. CP: culture-positive; CN: culture-negative; HC: healthy controls.

Fig. S6.

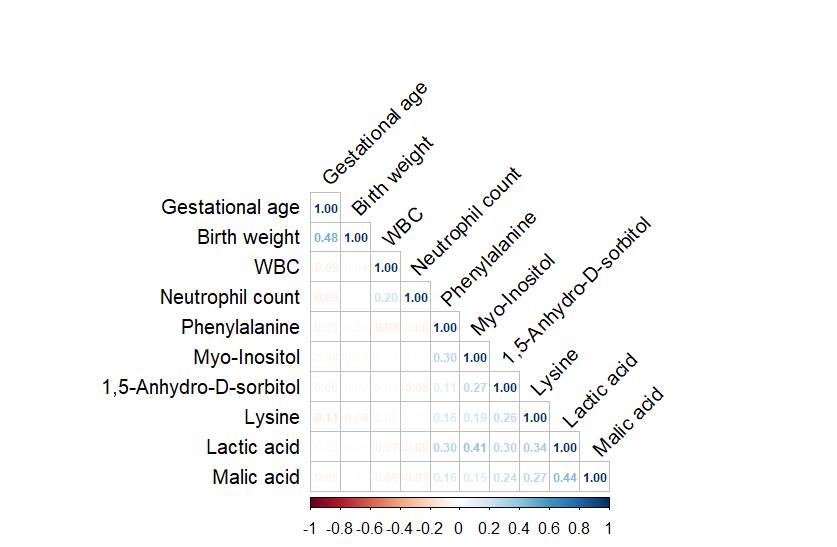

A

C

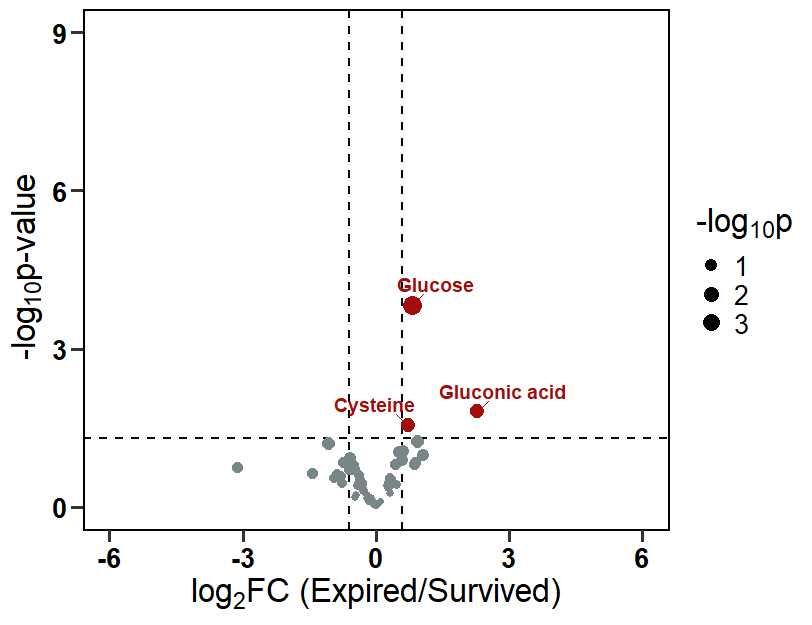

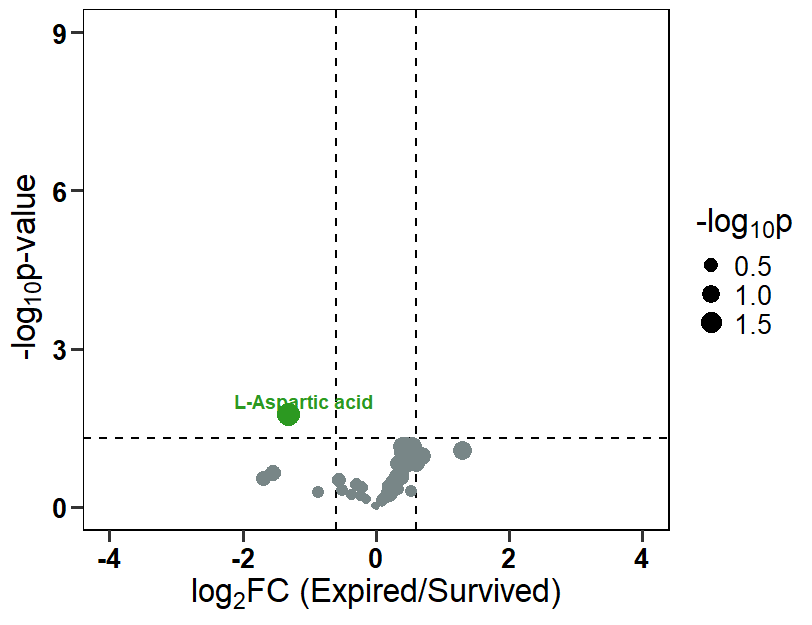

B

**Supplementary Figure S6. Correlation between sepsis serum metabolite signature and putative serum metabolite markers for disease severity. (A)** Correlation plot depicting the correlation between sepsis serum metabolite signature and gestational age, birth weight, WBC, and neutrophil count, quantified for sepsis diagnosis. **(B-C)** The volcano plot highlights the metabolites that differentiate expired neonates from surviving ones in the **(B)** CP and **(C)** CN groups. r: Pearson’s correlation coefficient; FC: Fold Change; WBC: White Blood Cells.

Fig. S7.

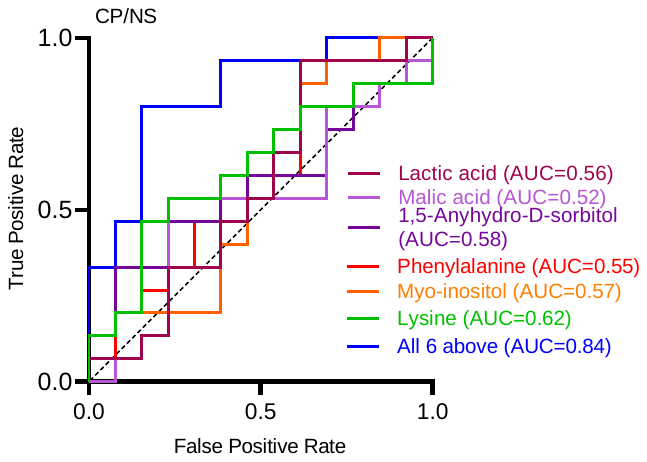

A

B

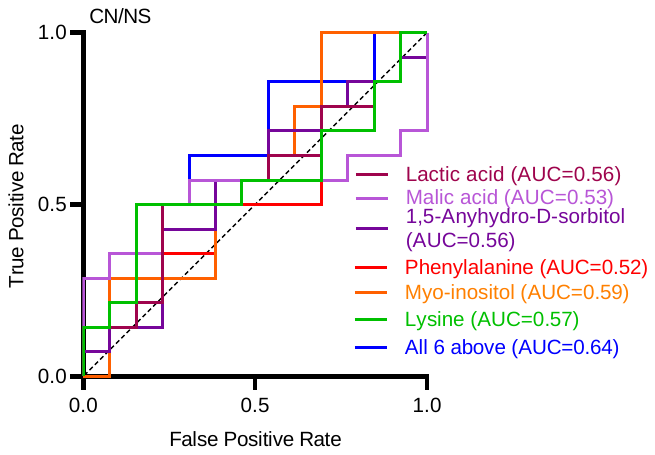

**Supplementary Figure S7. AUC of ROC of the biosignature consisting of 6 biomarkers depicts high predictive ability for CP and CN cases against the NS group (n=13-15 each).** AUC of ROC in **(A)** CP against NS group, and **(B)** CN against NS group. AUC: area under the curve; ROC: receiver operating curve; CP: culture-positive; NS: no sepsis; CN: culture-negative.

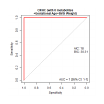

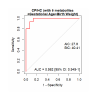

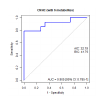

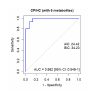
**Fig. S8.**

D

C

B

A

Supplementary Figure S8. AUC of ROC of serum metabolic biosignature considering their birth weight and gestational age. (A) AUC of ROC of metabolic signature differentiating CP from HC and (B) after normalizing with gestational age and birth weight. (C) AUC of ROC of metabolic signature differentiating CN and HC and (D) after normalizing data with gestational age and birth weight. ROC: Receiver Operating Curve; AUC: Area Under Curve; CP: Culture-positive; CN: Culture-negative; NS: No sepsis; HC: Healthy control.

Fig. S9.

B

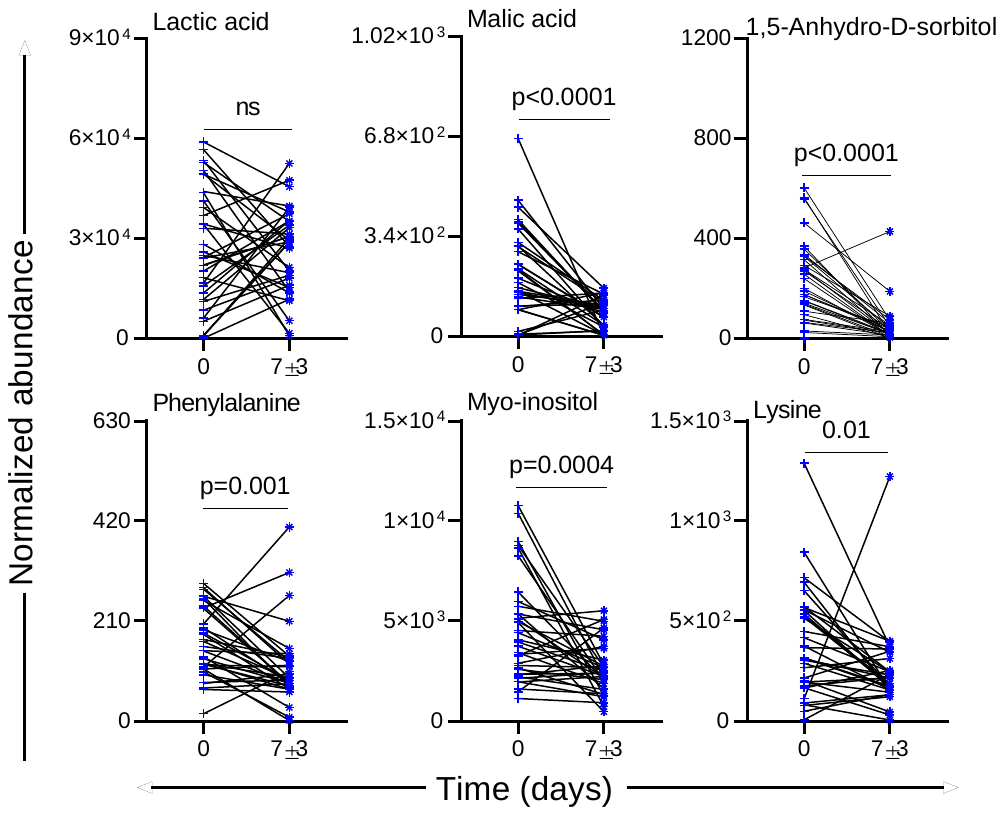

C

A

F

D

E

**Supplementary Figure S9. The trend in the abundance of deregulated metabolites in longitudinally followed-up NS cases upon recovery.** Dot plots depicting the trend of Lactic acid **(A)**, Malic acid **(B)**, 1,5-Anhydro-D-sorbitol **(C)**, Phenylalanine **(D)**, Myo-inositol **(E)**, and Lysine **(F)** in the NS group of an independent data set (n=36). NS: no sepsis.

**Fig. S10.**

**A**

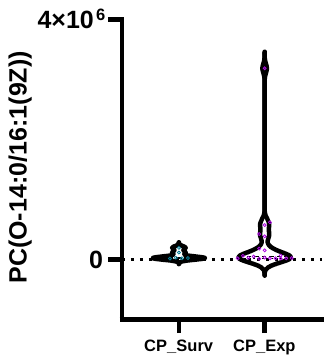

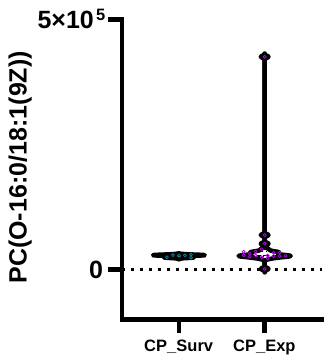

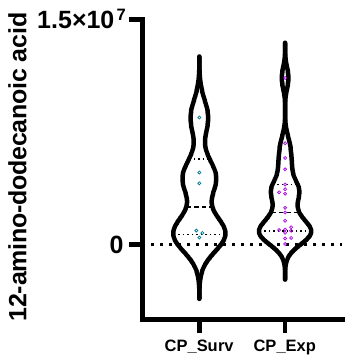

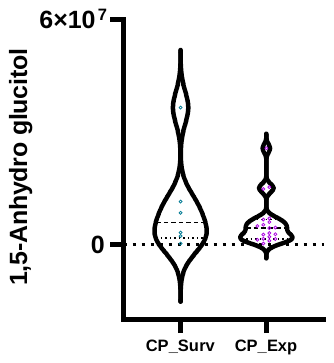

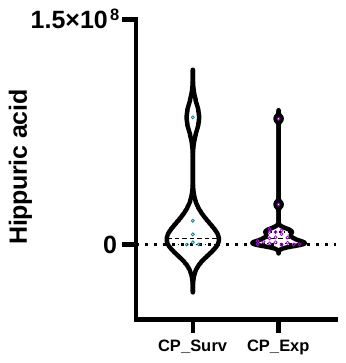

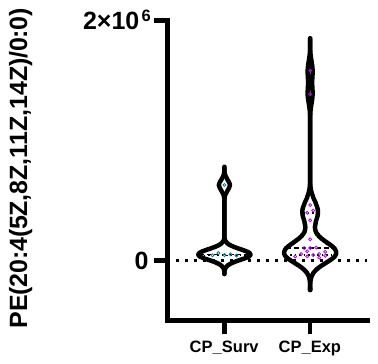

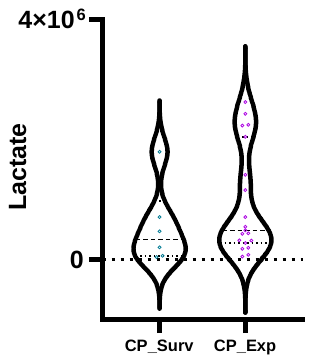

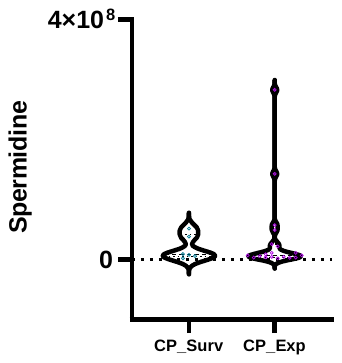

**B**

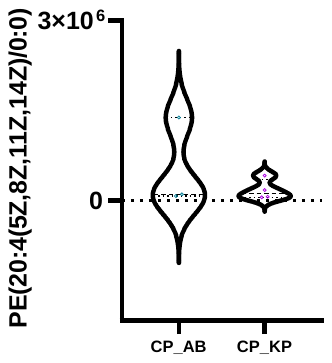

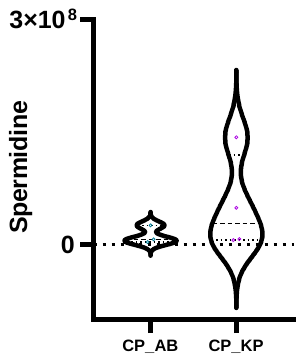

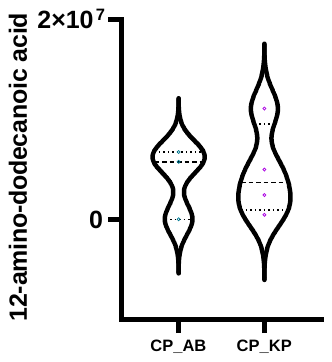

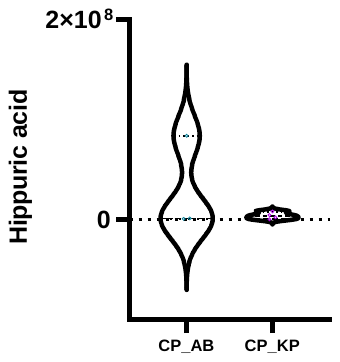

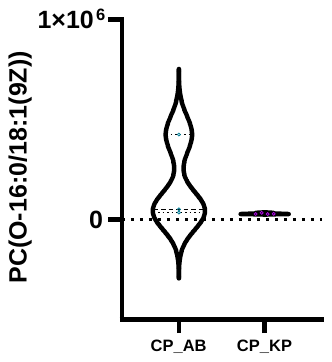

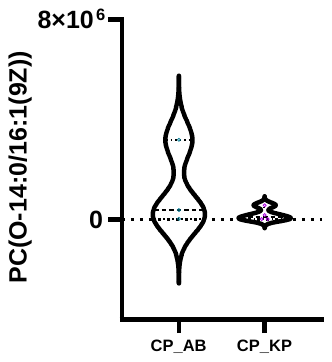

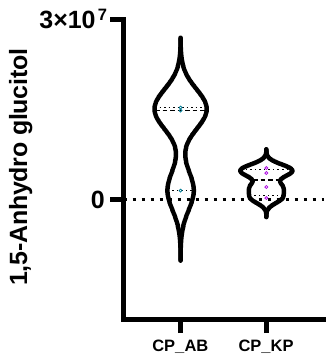

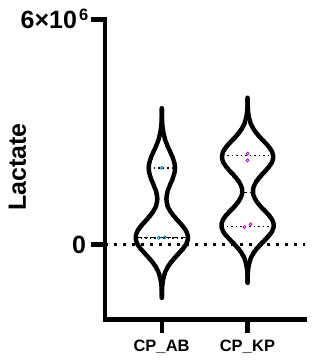

**Supplementary Figure S10. Deregulated serum metabolites and their distribution in culture-positive neonatal sepsis subjects in validation set-II.** Violin plots illustrating the abundance of identified metabolites comparing (A) survivors versus non-survivors among neonatal sepsis patients and between (B) *Acinetobacter baumannii*-positive (CP_AB) versus *Klebsiella pneumoniae*-positive (CP_KP) cases within the culture-positive neonatal sepsis subjects.

**Supplementary Table S1.**

**The Table containing individual patient details is presented in a separate Excel file.**

Supplementary Table S2. Group-specific epidemiological details and their significance in neonatal sepsis and control cases.

| **Clinical details** | **Total** | **Discovery set-I** | | | | | | | | | | **Discovery set-II** | | | | | | | | | |
| --- | --- | --- | --- | --- | --- | --- | --- | --- | --- | --- | --- | --- | --- | --- | --- | --- | --- | --- | --- | --- | --- |
|  |  | **Sub total** | **Sepsis** | | **Control** | | **p-value** | | | | | **Subtotal** | **Sepsis** | | **Control** | | **p- value** | | | | |
|  |  |  | **CP** | **CN** | **HC** | **NS** | **CP/HC** | **CN/HC** | **CP/CN** | **CP/NS** | **CN/NS** |  | **CP** | **CN** | **HC** | **NS** | **CP/HC** | **CN/HC** | **CP/CN** | **CP/NS** | **CN/NS** |
| **Number of neonates** | 600 | 71 | 16 | 24 | 14 | 17 |  |  |  |  |  | 269 | 63 | 69 | 69 | 68 |  |  |  |  |  |
| **Gender  (% female, total)** | 41.3,  248 | 42.3,  30 | 50,  8 | 41.7,  10 | 42.9,  6 | 35.3,  6 | 0.7 | 0.94 | 0.6 | 0.39 | 0.68 | 43.4,  117 | 31.7,  20 | 49.3,  34 | 50.7,  35 | 41.2,  28 | 0.02 | 0.86 | **0.04** | 0.23 | 0.38 |
| **Weight  (mean ± std. dev. in kg)** | 1.5 ±0.4 | 1.5 ±0.5 | 1.3 ±0.5 | 1.4 ±0.3 | 2 ±0.4 | 1.6 ±0.4 | **5E-04** | **1E-04** | 0.46 | 0.15 | 0.38 | 1.5 ±0.3 | 1.4 ±0.3 | 1.4 ±0.4 | 1.7 ±0.3 | 1.6  ±0.3 | **0.0001** | **3E-04** | 0.86 | **0.01** | **0.05** |
| **Mode of delivery  (% Caesarean, total)** | 46.3,  278 | 32.4,  23 | 31.3,  5 | 20.8,  5 | 50,  7 | 35.3,  6 | 0.3 | **0.05** | 0.26 | 0.81 | 0.31 | 57.6,  155 | 31.7,  20 | 40.6,  28 | 43.5,  30 | 51.5,  35 | 0.16 | 0.86 | 0.22 | **0.02** | 0.26 |
| **Gestation**  **(mean ± std. dev. in weeks)** | 31.5 ±1.9 | 31.4 ±1.9 | 30.9 ±1.7 | 30.8 ±2.1 | 32.9 ±1.5 | 31.7 ±1.6 | **0.002** | **0.007** | 0.48 | 0.16 | 0.24 | 31.4 ±1.7 | 30.9  ±1.9 | 30.9 ±1.8 | 32.3 ± 1.3 | 31.6 ±1.5 | **0.0001** | **1E-04** | 0.79 | **0.04** | **0.03** |
| **Outcome  (%, total)** | 23.7,  600 | 18.3,  71 | 43.8,  16 | 16.6,  24 | - | 11.8,  17 | - | - | 0.07 | 0.05 | 0.66 | 40.8,  250 | 77.3,  53 | 65,  63 | 1.5,  66 | 27.9,  68 | **0.0001** | **1E-04** | **0.15** | **0.001** | **1E-04** |
| **Difficulty breathing**  **(%, total)** | 19.1,  115 | 50,  52 | 43.8,  16 | 62.5,  22 | - | 33.3,  12 | - | - | 0.25 | 0.58 | 0.06 | 77.3,  181 | 81.1,  53 | 75.4,  61 | - | 76.1,  61 | 0.48 | 0.58 | 0.46 | 0.5 | 0.92 |
| **Apnea  (%, total)** | 15.8,  95 | 23.1,  52 | 18.8,  16 | 27.3,  22 | 50,  2 | 8.3, 12 | - | - | 0.54 | 0.45 | 0.22 | 6.5,  182 | 13.2,  53 | 3.2,  63 | - | 4.5,  66 | 0.37 | 0.12 | **0.06** | 0.1 | 0.68 |
| **WBC count**  **(mean ± std. dev. in 10^3^/μL)** | - | - | 18.8 ±18 | 24.2 ±21.2 | - | 24.1 ±14.1 | - | - | 0.59 | 0.08 | 0.31 | - | 11.2  ±9.8 | 16.9 ±17.8 | - | 13.4 ±9.4 | - | - | **0.03** | **0.03** | 0.8 |
| **Absolute neutrophil count**  **(mean ± std. dev. in 10^3^)** | - | - | 3.8 ±2.5 | 3.7 ±2.5 | - | 6.9 ±3.4 | - | - | 0.97 | **0.03** | **0.03** | - | 4.8 ±3.1 | 4.2 ±3.8 | - | 4.1 ±2.0 | - | - | 0.34 | 0.71 | 0.25 |

| **Clinical details** | **Validation set-I** | | | | | | | | | | **Follow-up set** | | | | | | | **Validation set-II (LC-MS)** | | | | | | | | |
| --- | --- | --- | --- | --- | --- | --- | --- | --- | --- | --- | --- | --- | --- | --- | --- | --- | --- | --- | --- | --- | --- | --- | --- | --- | --- | --- |
|  | **Subtotal** | **Sepsis** | | **Control** | | **p-value** | | | | | **Sub**  **total** | **Sepsis** | **Control** | | **p-value** | | | **Sub**  **total** | **Sepsis** | | **Control** | | **p-value** | | | |
|  |  | **CP** | **CN** | **HC** | **NS** | **CP/HC** | **CN/HC** | **CP/CN** | **CP/NS** | **CN/NS** |  | **CP** | **CN** | **NS** | **CP/CN** | **CP/NS** | **CN/NS** |  | **CP** | **CN** | **HC** | **NS** | **CP/HC** | **CN/HC** | **CP/CN** | **CP/NS** |
| **Number of neonates** | 60 | 15 | 15 | 15 | 15 |  |  |  |  |  | 100 | 29 | 35 | 36 |  |  |  | 100 | 25 | 25 | 25 | 25 |  |  |  |  |
| **Gender  (% female, total)** | 40,  24 | 60,  9 | 46.7,  7 | 20,  3 | 33.3,  5 | 0.7 | 0.94 | 0.6 | 0.27 | 0.46 | 46,  46 | 48.3,  14 | 40,  14 | 50,  18 | 0.51 | 0.89 | 0.4 | 42,  42 | 44,  11 | 40,  10 | 44,  11 | 40,  10 | 1.0 | 0.7 | 0.7 | 0.7 |
| **Weight**  **(mean ± std. dev. in kg)** | 1.5 ±0.4 | 1.5 ±0.4 | 1.3 ±0.3 | 1.7 ±0.3 | 1.7 ±0.5 | 0.37 | **0.004** | 0.15 | 0.29 | **0.01** | 1.5 ±0.3 | 1.5 ±0.3 | 1.5 ±0.3 | 1.6 ±0.4 | 0.77 | 0.1 | 0.1 | 1.5±0.3 | 1.4 ± 0.2 | 1.4 ± 0.3 | 1.6 ± 0.3 | 1.6 ± 0.3 | 0.007 | 0.02 | 1.0 | 0.007 |
| **Mode of delivery  (% Caesarean, total)** | 28.3,  17 | 40,  6 | 13.3,  2 | 26.7,  84 | 33.3,  5 | 0.44 | 0.37 | 0.11 | 0.71 | 0.21 | 46,  46 | 48.3,  14 | 51.4,  18 | 38.9,  14 | 0.8 | 0.45 | 0.29 | 37, 37 | 48, 25 | 32, 25 | 28,25 | 40, 25 | 0.1 | 0.7 | 0.2 | 0.5 |
| **Gestation**  **(mean ± std. dev. in weeks)** | 31.6 ±1.9 | 30.9 ±2 | 30.5 ±1.9 | 32.6 ±1.2 | 32.3 ±1.5 | **0.01** | **0.004** | 0.5 | **0.04** | **0.01** | 31.5 ±1.8 | 31.2 ±2 | 31.2 ±1.7 | 32 ±1.6 | 0.88 | **0.12** | **0.04** | 31.6 ± 1.7 | 31.1 ± 1.9 | 31.1 ± 1.5 | 32.4 ± 1.2 | 31.8 ± 1.6 | 0.005 | 0.001 | 1.0 | 0.1 |
| **Outcome  (%, total)** | 30,  60 | 60,  15 | 60,  15 | 0 | 20,  15 | - | - | 1 | **0.03** | **0.03** | - | - | - | - | - | - | - | 43,  95 | 76, 25 | 63.6, 22 | - | 34.7, 23 | - | - | 0.3 | 0.004 |
| **Difficulty breathing  (%, total)** | 85,  40 | 100,  12 | 100,  13 | - | 33.3,  12 | - | - | 1 | **0.01** | **0.01** | 84,  99 | 86.2,  29 | 88.6,  35 | 80,  35 | 0.78 | 0.51 | 0.33 | 68,  95 | 100, 25 | 90.9, 22 | - | 86.9, 23 | - | - | 0.1 | 0.06 |
| **Apnea  (%, total)** | 12.5,  40 | 25,  12 | 7.7,  13 | - | 6.7,  15 | - | - | 0.63 | 0.21 | 0.92 | 11,  100 | 20.7,  29 | 11.4,  35 | 2.8,  35 | 0.32 | **0.05** | 0.19 | 3, 97 | 4, 25 | 9.0, 22 | - | - | - | - | 0.4 | - |
| **WBC count**  **(mean ± std. dev. in 10^3^/μL)** | - | 10.1 ±5 | 18.2 ±12.6 | - | 13.5 ±5.8 | - | - | 0.08 | 0.25 | 0.35 | 13.3 ±8.9 | 11.4 ±9 | 13.5 ±10.8 | 14.7 ±5.6 | 0.56 | **0.005** | **0.02** | - | 12.5 ± 10.6 | 8.4 ± 7.1 | - | 11.1 ± 7.8 | - | - | 0.2 | 0.9 |
| **Absolute neutrophil count  (mean ± std. dev. in 10^3^)** | - | 3.6 ±2.28 | 7.9 ±9.8 | - | 5.9 ±3.6 | - | - | 0.22 | 0.16 | 0.97 | 5.7 ±5.4 | 3.6 ±2.6 | 7.1 ±7.4 | 5.7 ±2.7 | **0.04** | **0.007** | 0.67 | - | 0.9 ± 2.7 | 0.8 ± 2.1 | - | 1.7 ± 3.2 | - | - | 0.06 | 0.1 |

**Supplementary Table S3**. Distribution of *Acinetobacter baumannii* and *Klebsiella pneumoniae* cases in the culture-positive neonatal sepsis cases.

| Pathogen | Discovery set-I | Discovery set-II | Validation set-I | Follow up  set | Validation set-II |
| --- | --- | --- | --- | --- | --- |
| CP sepsis cases | 16 | 63 | 15 | 29 | 25 |
| *Acinetobacter baumannii* | 6 | 13 | 6 | 4 | 3 |
| *Klebsiella pneumonia* | - | 19 | 1 | 6 | 4 |
| *CoNS* | 3 | 5 | 1 | - | - |

**Supplementary Table S4**. Study group specific distribution of gestational age and birth weight of neonatal sepsis cases (CP/CN: culture +/-ve) and controls (NS/HC: no-sepsis and healthy controls).

| Study groups | Gestational age  (weeks) | Birth weight  (kg) |
| --- | --- | --- |
| Discovery set-I | | |
| CP | 30.9±1.7 | 1.3±0.5 |
| CN | 30.8±2.1 | 1.4±0.3 |
| NS | 31.7±1.6 | 1.6±0.4 |
| HC | 32.9±1.5 | 2.0±0.4 |
| Discovery set-II | | |
| CP | 30.9±1.9 | 1.4±0.3 |
| CN | 30.9±1.2 | 1.5±0.4 |
| NS | 31.6±1.5 | 1.6±0.3 |
| HC | 32.3±1.3 | 1.7±0.3 |
| Validation set-I | | |
| CP | 30.9±2.0 | 1.5±0.4 |
| CN | 30.5±1.9 | 1.3±0.3 |
| NS | 32.6±1.5 | 1.7±0.5 |
| HC | 32.3±1.5 | 1.7±0.3 |
| Follow up set | | |
| CP | 31.2±2.0 | 1.5±0.3 |
| CN | 31.2±1.7 | 1.5±0.3 |
| NS | 32.0±1.6 | 1.6±0.4 |
| Validation set-II | | |
| CP | 31.1 ± 1.9 | 1.4 ± 0.2 |
| CN | 31.1 ± 1.5 | 1.4 ± 0.3 |
| NS | 31.8 ± 1.6 | 1.6 ± 0.3 |
| HC | 32.4 ± 1.2 | 1.6 ± 0.3 |

**Supplementary Table S5.** Fold change (FC) and (p-value) of deregulated serum metabolites that could differentiate sepsis from control groups.

| **Metabolic**  **feature** | **Discovery set-I** | | | | | **Discovery set-II** | | | | | **Validation set** | | | | |
| --- | --- | --- | --- | --- | --- | --- | --- | --- | --- | --- | --- | --- | --- | --- | --- |
|  | **CP/HC** | **CN/HC** | **CP/CN** | **CP/NS** | **CN/NS** | **CP/HC** | **CN/HC** | **CP/CN** | **CP/NS** | **CN/NS** | **CP/HC** | **CN/HC** | **CP/CN** | **CP/NS** | CN/NS |
|  | **log_2_FC (p-value)** | | | | | | | | | | | | | | |
| 1,5-Anhydro-D-sorbitol | 0.04 (0.9) | 0.8 (0.02) | -0.7 (0.01) | -0.08 (0.8) | 0.6 (0.04) | 1.6 (<0.0001) | 1.9 (<0.0001) | -0.3 (0.1) | -0.3 (0.1) | 0.1 (0.0004) | 2.7 (0.0002) | 2.3 (0.003) | 0.4 (0.3) | 0.3 (0.3) | -0.07 (0.9) |
| Lactic acid |  |  |  |  |  | 1 (<0.0001) | 0.9 (<0.0001) | 0.1 (0.5) | 0.1 (0.5) | 1 (0.002) | 1 (0.01) | 1 (0.05) | -0.01 (1) | 0.3 (0.5) | 0.3 (0.6) |
| Lysine | 0.8 (0.04) | 0.6 (0.05) | 0.2 (0.5) | 0.4 (0.2) | 0.2 (0.4) | 0.6  (0.01) | 1.3 (<0.0001) | -0.7 (0.01) | -0.6 (0.01) | 0.02 (0.9) | 1.6 (0.01) | 1 (0.07) | 0.6 (0.2) | 0.6 (0.2) | -0.02 (1) |
| Malic acid |  |  |  |  |  | 0.9  (0.02) | 0.9 (0.003) | 0.01 (1) | -0.05 (0.9) | -0.06 (0.8) | 0.9 (0.0008) | 1.4 (0.03) | -0.6 (0.2) | -0.06 (0.8) | 0.5 (0.3) |
| Myo-inositol | 0.8 (0.01) | 1  (0.001) | -0.3 (0.2) | 0.1 (0.6) | 0.5 (0.07) | 1 (<0.0001) | 1 (<0.0001) | 0.03 (0.9) | -0.03 (0.9) | -0.06 (0.7) | 0.9 (0.01) | 1 (0.01) | -0.08 (0.8) | -0.3 (0.3) | -0.2 (0.5) |
| Phenylalanine | 1 (0.02) | 0.4 (0.2) | 0.6 (0.1) | 0.4 (0.2) | -0.1 (0.6) | 1.2 (0.03) | 0.8 (<0.0001) | 0.5 (0.3) | 0.5 (0.3) | 0.02 (0.9) | 0.9 (0.01) | 0.6 (0.1) | 0.3 (0.3) | 0.3 (0.4) | -0.08 (0.8) |

**Supplementary Table S6.** List of the serum metabolites selected for biomarker identification and validation.

| **Discover set-I (n=52)** | **Discovery set-II,**  **Validation set (n=56)** |
| --- | --- |
| 1,5-Anhydro-D-sorbitol | 1,2-Propanediol |
| 17a-Hydroxyprogesterone | 1,5-Anhydro-D-sorbitol |
| 19-Norethindrone | 2,3,4-Trihydroxybutyric acid |
| 2,3,4-Trihydroxybutyric acid | 2-Hydroxybutyric acid |
| 3,4,5-Trihydroxypentanoic acid | 3-Aminoisobutyric acid |
| Aconitic acid | 3-Hydroxybutyric acid |
| Alanine | Alanine |
| Allose | Allonic acid |
| Androst-2-en-17-amine | Aminomethane |
| Arabinose | Arabinose |
| Arachidonic acid | Arabitol |
| Asparagine | Butanoic acid |
| Benzoic acid | Cadaverine |
| Beta-Gentiobiose | Citric acid |
| Butanoic acid | Creatinine |
| Cholesterol | Cystein |
| Citric acid | D-Gluconic acid |
| Cysteine | Ethanolamine |
| D-Erythro-Pentofuranose | Galactose |
| d-Galactose | Gluconic acid lactone |
| Diethyl phthalate | Glucose |
| Diethyl tartrate | Glutamic acid |
| Fructose | Glycine |
| Galactitol | Hippuric acid |
| Glucose | Hydrocinnamic acid |
| Glutamic acid | Hydroxyproline |
| Glycine | Indole pyruvic acid |
| Histidine | Indole-3-acetic acid |
| Hydroxyproline | Isoleucine |
| Isoleucine | Lactic acid |
| Linoleic acid | Aspartic acid |
| Methionine | Leucine |
| Lysine | Lysine |
| Maltose | Lyxofuranose |
| Mannitol | Malic acid |
| Myo-Inositol | Mannose |
| Oleic acid | Methionine |
| Ornithine | Myo-Inositol |
| Oxoproline | N,N-Dimethylglycine |
| Phenylalanine | Oleic acid |
| Phosphoric acid | Ornithine |
| Pregnan-18-oic acid | Oxalic acid |
| Pseudouridine | Phenylalanine |
| Serine | Phosphoric acid |
| Succinic acid | Proline |
| Talose | Propanoic acid |
| Tartaric acid | Serine |
| Threonine | Sorbitol |
| Tryptophan | Tagatose |
| Tyrosine | Threonine |
| Urea | Tyrosine |
| Uridine | Unknown 1 |
| Valine | Unknown 2 |
|  | Valine |
|  | Xylo-hexos-5-ulose |
|  | Xylose |

**Supplementary Table S7**. The criteria used for neonatal sepsis suspicion, confirmation and grouping are adopted in this study.

|  | **Clinical criteria for suspicion of sepsis (any one or more)** | **Laboratory criteria for confirmed sepsis (any two parameters are considered for positive screening)** |
| --- | --- | --- |
| 1 | Difficulty feeding | White blood cell (WBC) count (absolute numbers/ microlitre) <4.0 x 109 cells/L |
| 2 | Convulsions or abnormal posturing | Absolute neutrophil count < 1.5 x 109 cells/L |
| 3 | No movement or movement only when stimulated OR significant lethargy | C-reactive protein (CRP) absolute value in mg/L > 6mg/L |
| 4 | Fever | Total leucocyte count (TLC) < 5000/cmm |
| 5 | Cold to touch | Band to total plymorph ratio >2 |
| 6 | New onset or increased apnea | Micro ESR > 15 mm in 1^st^ hour |
| 7 | Bleeding from the gastrointestinal tract |  |
| 8 | Hypotonia or floppiness |  |
| 9 | Difficulty breathing |  |
| 10 | Bloody/bilious vomiting |  |
| 11 | Diarrhea (watery stool) |  |
| 12 | Pus from the umbilical stump |  |
| 13 | Discharge from ear |  |
| 14 | Mottling |  |
| 15 | Bleeding from skin/mucosa/lungs |  |
| 16 | Others (Specify) |  |
| 17 | Temperature >37.5°C |  |
| 18 | Heart Rate > 180/min |  |
| 19 | Capillary refill time (CRT) > 3 sec |  |
| 20 | Respiratory rate > 60/min |  |
| 21 | Severe chest in-drawing |  |
| 22 | Movement only when stimulated |  |
| 23 | Cyanosis |  |
| 24 | Sclerema |  |
| 25 | Temperature <36.5°C |  |
| 26 | Heart Rate < 100/min |  |
| 27 | Grunting |  |
| 28 | Bulging fontanelle |  |
| 29 | Abdominal distension |  |
| 30 | Abdominal erythema |  |
| 31 | Abdominal tenderness |  |
| 32 | Meningitis |  |
| 33 | PNEUMONIA |  |
| 34 | Peritonitis |  |
| 35 | Necrotizing enterocolitis |  |

**Supplementary Table S8.** Definition used for classification of the neonatal sepsis suspected cases.

| **Diagnosis** | **Working definitions** |
| --- | --- |
| **Culture positive sepsis**  **(CP)** | Baby unwell/maternal-perinatal risk factors **AND** must meet **ANY ONE** of the following criteria  1. True pathogen detected: Baby has a recognized pathogen cultured from 1 or more blood cultures and organism cultured is not related to an infection at another site 2. CoNS: CoNS is cultured from 1 or more blood samples drawn on separate occasions and organism cultured is not related to an infection at another site AND Physician institutes appropriate treatment OR death/LAMA/referred before completing appropriate therapy. |
| **Culture negative sepsis**  **(CN)** | Baby unwell/maternal-perinatal risk factors **AND** ***all*** of the following criteria:  1. Blood culture not done or no organisms detected in blood. 2. Positive septic screen OR physician institutes appropriate treatment for sepsis OR death/LAMA/referred before completing appropriate treatment. |
| **No sepsis**  **(NS)** | Baby unwell/maternal-perinatal risk factors **AND** following criteria:  1. Negative septic screen and blood culture  2. Physician did not treat/stopped treatment before 5 days as it was not considered to be sepsis |
